# Predicting cognitive decline in a low-dimensional representation of brain morphology

**DOI:** 10.1101/2022.11.23.22282135

**Authors:** Rémi Lamontagne-Caron, Patrick Desrosiers, Olivier Potvin, Simon Duchesne, Nicolas Doyon

## Abstract

Identifying early signs of neurodegeneration due to Alzheimer’s disease (AD) is a necessary first step towards preventing cognitive decline. Individual cortical thickness measures, available after processing anatomical magnetic resonance imaging (MRI), are sensitive markers of neurodegeneration. However, normal aging cortical decline and high inter-individual variability complicate the comparison and statistical determination of the impact of AD-related neurodegeneration on trajectories. In this paper, we computed trajectories in a 2D representation of a 62-dimensional manifold of individual cortical thickness measures. To compute this representation, we used a novel, nonlinear dimension reduction algorithm called Uniform Manifold Approximation and Projection (UMAP). We trained two embeddings, one on cortical thickness measurements of 6,237 cognitively healthy participants aged 18 to 100 years old and the other on 233 mild cognitively impaired (MCI) and AD participants from the longitudinal database, the Alzheimer’s Disease Neuroimaging Initiative database (ADNI). Each participant had multiple visits (*n* ≥ 2), one year apart. The first embedding’s principal axis was shown to be positively associated (*r* = 0.65) with participants’ age. Data from ADNI is projected into these 2D spaces. After clustering the data, average trajectories between clusters were shown to be significantly different between MCI and AD subjects. Moreover, some clusters and trajectories between clusters were more prone to host AD subjects. This study was able to differentiate AD and MCI subjects based on their trajectory in a 2D space with an AUC of 0.80 with 10-fold cross-validation.

## Introduction

A proper understanding of brain morphology trajectory during cognitively healthy aging can be leveraged to detect departures due to neurodegeneration (e.g. from Alzheimer’s disease (AD)) and therefore serve as an early indicator of incipient dementia. As the socio-economic costs of AD increase with the aging of global populations^1,2^, early detection becomes the only approach which allows for the possibility of interventions able to reduce the risk of conversion to dementia.

Brain morphology can be assessed via magnetic resonance imaging (MRI)^3–6^. Morphometry has been shown to be a sensitive marker of neurodegeneration^7,8^. However, normal aging cortical decline and high inter-individual variability complicate the comparison and statistical determination of the impact of AD-related neurodegeneration on trajectories. Further, the high-dimensional nature of morphometry (whereas the available measurements routinely surpass the number of study participants) necessitate the use of a transformation into lower-dimensional spaces in order to perform statistical analyses, modeling or machine learning^9–12^. In these resulting spaces of low dimensionality, the task then turns to the identification of criteria with which to determine whether a given brain morphology trajectory is pathological or not, which implies having comparative instances of both successful and pathological natures.

High-to-low dimensionality transformations have been carried out via a number of approaches in the past. In some instances, dimensionality reduction or machine learning algorithms were applied directly to the raw voxel data^13–17^. Other approaches first use tools (e.g. FreeSurfer^18^) to extract features such as volumes, thicknesses or surface areas for individual brain structure, then perform dimensionality reduction on the feature vectors^19–24^. While the first family of approaches may yield classifiers with adequate accuracy, the lower-dimensional space and resulting classification functions are usually difficult to interpret. On the other hand, the second family of approaches allows the association between principal dimensions to interpretable features with biological meaning^25^, which in turn might suggest intervention strategies. In recent years, the brain age field of research has used both approaches to predict biological age for the whole spectrum of cognitive health with successful results. In turn, this technique has also shown potential for the prediction of cognitive decline^26–30^.

There remains a question as to whether linear or nonlinear dimensionality reduction techniques should be utilized. In our context, studies of brain morphology in aging have concluded that whole brain volume trajectories as well as microstructural and focal markers exhibit nonlinear patterns^31,32^. More recently, we have shown that atrophy across the brain follows for the most part a quadratic trajectory^33^. Thus, in order for any lower-dimensional embedding to correctly encompass this multi-dimensional problem, a nonlinear dimension reduction algorithm has to be used. Utilizing a non-linear technique removes the need to have a linear combination as a prior for the representation in a low-dimensional embedding.

In that class, Uniform Manifold Approximation and Projection (UMAP) is a novel entry, first published in 2018.^34^ UMAP’s backbone rests on Riemannian manifold theory and topological data analysis. Otherwise, its main advantage over other nonlinear methods, like t-distributed Stochastic Neighbor Embedding (t-SNE)^35^ or nonlinear principal component analysis^36^, comes from its agility, performance and conservation of global structures. Arising from Riemannian mathematics, the algorithm is highly generalizable and doesn’t necessitate strict priors on the data, such as the features being linearly dependent between each other. As to not imbue priors to the low-dimension embedding, UMAP is a good candidate. Moreover, the algorithm works on two main hyperparameters : `n_neighbors` and `min_dist`. The choice of `n_neighbors` selects how much the local structures are expressed in the low-dimension embedding, which in our case allows us to chose the optimal embedding that works best for the problem. `min_dist` is the minimum distance between datum in low-dimension i.e. two identical feature vectors will occupy the same point after UMAP, if the distance parameter is set to zero. In our case, it can make sure the trajectories in low-dimensions will be truly related to their high-dimension sibling which is essential.^34^ Further, as opposed to highly nonlinear deep learning networks, a UMAP based method is interpretable through visualization. It is therefore a suitable technique to handle the nonlinear nature of brain morphological trajectories and embed our data vectors in a lower-dimensional space. Examples of similar applications can be found in Campbell et al. and Becht et al.^37,38^

In this article, we present our work on brain morphology trajectory estimation on a number of well-known, publicly available datasets composed of individuals across the spectrum of cognitive health. Our general goal is to use UMAP to generate a reference low-dimensional representation of morphology from vectors of features describing the thicknesses of different brain regions; then project in this space data from individuals across the spectrum of cognitive decline. A discretization scheme alongside probabilistic analysis of trajectories is used to achieve the best possible accuracy at predicting the longitudinal cognitive outcome in individuals. As a first experiment, we built the reference space using a dataset composed of cognitively healthy (CH) individuals, in which we projected data on subjects with mild cognitive impairment (MCI; a prodromal state of AD) that either progressed to dementia due to probable Alzheimer’s disease (MCI-AD) or remained cognitively stable (MCI-MCI). In a second experiment, we used data on MCI-AD and MCI-MCI individuals to form the reference space as a basis for classification. In both experiments we tested the association with aging.

## Methods

Figure 1 shows a diagram of the data processing pipeline used for this research, described in more detail in the following section.

**Figure 1.**
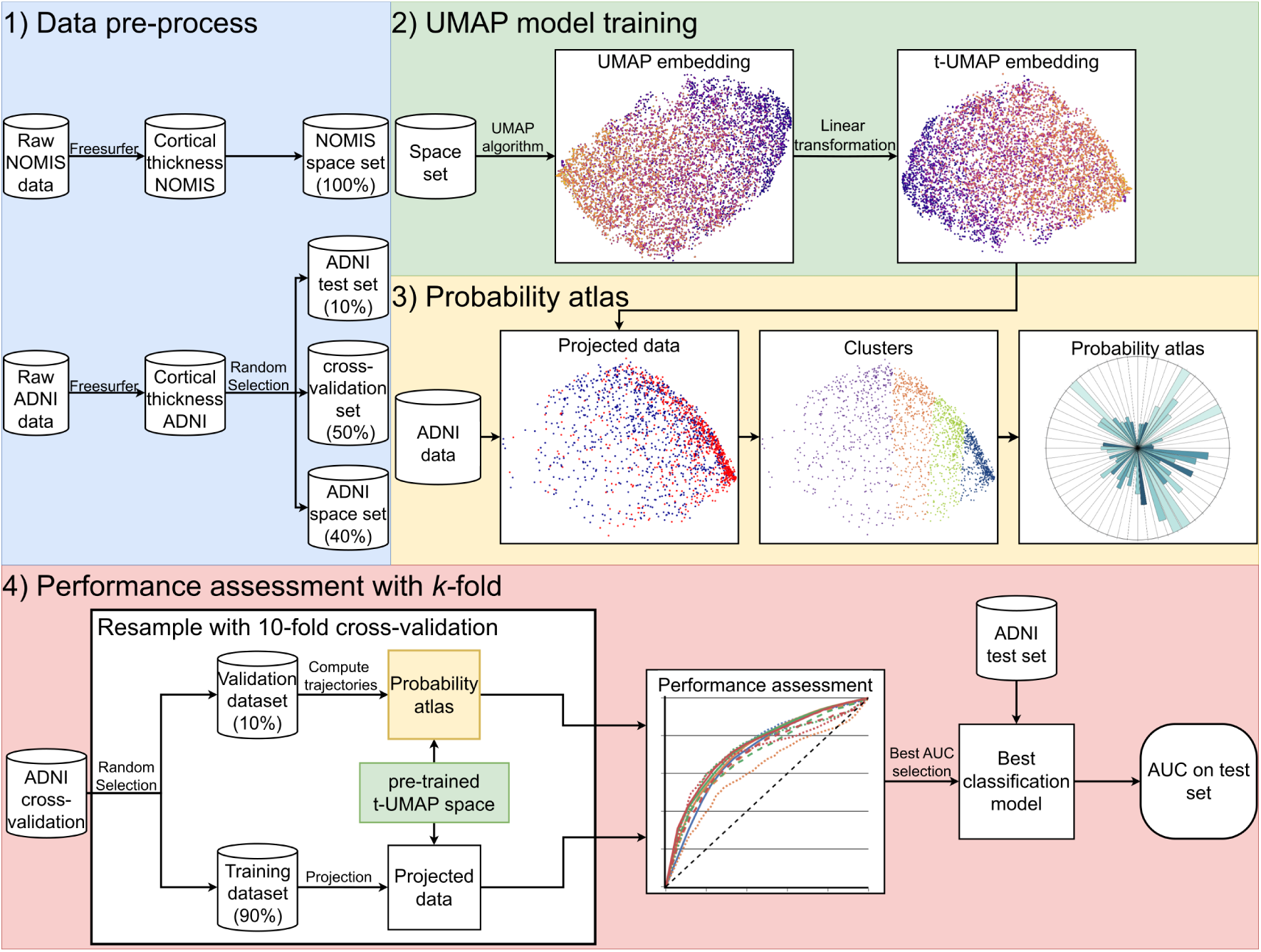
Methodological pipeline for the prediction of cognitive outcome. **1)** We separate the different datasets (NOMIS and ADNI) into the necessary subsets (test, cross-validation and space set). **2)** Different UMAP models are trained on one of the two space sets which results in the an embedding referred to as “UMAP embedding”. This embedding is linearly transformed so its axes are align with its principal axes. This transformation in turns results in a new embedding referred to as “t-UMAP embedding”. The latter embedding is fed to the probability atlas algorithm. The figures displayed under “UMAP embedding” and “t-UMAP embedding” were made using the NOMIS dataset with `n_neighbors` = 20 as the UMAP parameter. **3)** The probability atlas algorithm reduces the longitudinal data (ADNI for this paper) using the previously trained embedding, clusters the resulting data and computes the population and probability associated with each trajectory. **4)** Finally, the performance is assessed through 10-fold cross-validation using the “cross-validation set”, and the model with the best AUC is tested on the test set to generate the final accuracy of the method.

### Ethics

Approval for the study was obtained from the Ethics review committee of the CERVO Brain Research Center (NSM-2021-2022). Approval for this study was obtain from the local ethics board and informed consent for all participants were obtained for all datasets in this study (see Acknowledgements). All methods were carried out in accordance with relevant guidelines and regulations.

### Participants

#### NOMIS

The NOMIS database^33^ was used to train UMAP’s dimensionality reduction model. The database is composed of a total of 6,237 CH participants (3,556 women), aged 18 to 100 years old (median 56 y.o.), and aggregated from 26 different databases^33^. Participants had no history of: psychotic disorders, mood, anxiety, post-traumatic stress, or substance abuse/dependence disorders; neurodegenerative and neurological disorders; head injury with loss of consciousness/amnesia; lead poisoning; or geriatric depression. Since those participants were scanned once, the NOMIS data is cross-sectional.

#### ADNI

The Alzheimer’s Disease Neuroimaging Initiative (ADNI) database (ADNI1, GO, 2 and 3)^39^ was used to test the UMAP model for the prediction of cognitive decline. Launched in 2003 as a public-private partnership, led by Principal Investigator Michael W. Weiner, MD, ADNI’s goal has been to test whether serial MRI, positron emission tomography, other biological markers, and clinical and neuropsychological assessment can be combined to measure the progression of MCI and early AD. For up-to-date information, see www.adni-info.org. All ADNI participants with longitudinal time points and at least one follow-up after the baseline scan were selected for this project, for a total of 1354 participants (604 women). We used ADNI’s clinical diagnostic categorization at baseline (526 CH; 828 MCI; 0 AD) and last available follow-up (477 CH, 537 MCI; 340 AD), see Supplementary Table S5 for more details.

#### Diagnostic criteria

For the first ADNI cohort, the criteria for classification between CH, MCI and AD were based on memory loss, the Mini-Mental State Examination (MMSE), the Clinical Dementia Rating score and one paragraph from the Logical Memory II subscale of the Wechsler Memory Scale–Revised. For memory loss, participants needed to have complained about lapses in their memory to be classified as MCI or AD, while CH subjects did not experience memory lapses. With the MMSE, CH and MCI subjects had to have a score between 24 and 30, and for AD, 20-26. Cognitively healthy participants had to have a Clinical Dementia Rating score of 0, MCI 0.5 with the memory box score being 0.5 and AD a score of 0.5 or 1. For the last test, result depended on education. CH participants had a score ≥ 9 for 16 years of education, ≥ 5 for 8 to 15 years of education and ≥ 3 for 0-7 years of education. In the other hand, MCI and AD subjects had to have a score ≤ 8, ≤ 4 and ≤ 2 for the same education groups^40^.

#### Outcome

Throughout this article, we will refer to the evolution of clinical diagnostic categories as trajectory types. For instance, a subject with a MCI diagnostic at their first and last MRI was considered “stable” (MCI-MCI) while another who developed AD at a later stage would be a “converter” (MCI-AD).

### MRI data

#### NOMIS

Each NOMIS participant had an available T1-weighted MRI. Because the origin of the data is multiple, the protocol, MRI manufacturer and magnetic field strengths used were not uniform throughout the sample. Regarding manufacturers, 1,160 scans (18.60%) were made with a Philips Medical Systems MRI (Best, Netherlands), 3,853 (61.77%) with a Siemens Healthcare (Erlangen, Germany) scanner and 1,224 (19.63%) using GE Healthcare’s products (Milwaukee, WI). Of these scans, 4,778 were done at magnetic field strengths of 3.0 Tesla and 1,459 at a field of 1.5 Tesla.

Part of the data used in the preparation of this article were obtained from the [Parkinson’s Progression Markers Initiative (PPMI) database](https://www.ppmi-info.org/access-data-specimens/download-data). For up-to-date information on the study, visit www.ppmi-info.org.

#### ADNI

Participants were scanned on different MRI units albeit with a standardized protocol^41^. In total, there were 7414 scans and 4157 of those from MCI individuals at baseline. From those, the distribution of scans was similar to NOMIS with 547 (13.16%) using Philips Healthcare products, 2048 (49.26%) on Siemens Medical Systems units and 1562 (37.58%) on GE Healthcare. MRIs were taken with two field strengths; 2157 at 3.0 Tesla and 2000 at 1.5 Tesla. The age distribution was similar between MCI-AD (range: 55.1-95.9 y/o; average 76.1, SD 7.2 y/o) and MCI-MCI indviduals (range: 55.2-97.4 y/o; average 74.6, SD 7.7 y/o). For a full list of ADNI’s subjects, refer to Supplementary Table S6 to S10.

#### ADNI pre-processing

ADNI being a longitudinal, multi-centric study, some subjects were scanned every six months while others every year, with different lengths of follow-up (median of 10.2 months, mean of 11.04 months and a standard deviation of 7.8 months). Thus, before being processed, ADNI longitudinal data was binned into standardized time step of 1 year (+/- 5 months), based on the distribution of time steps in the index. This process and its impact on the data is documented in Supplementary Figures S1 to S4. This served to make trajectories comparable among participants. Further, in ADNI1/GO, some participants were scanned initially at a field of 1.5T, but at every other follow-up at 3.0 T. In these cases the first scan was discarded in order to not bias morphometry estimates by scanner and field strength changes. In fine, the ADNI sample consisted of 1780 scans from 549 MCI subjects with at least one follow-up, an average age of 74.6 (7.8) y/o for the MCI-MCI and 76.8 (7.3) y/o for MCI-AD. The distribution of regularized time steps had a median of 1.01, a mean of 1.00 and a standard deviation of 0.14.

#### Image processing

Each T1-weighted MRI was processed using *FreeSurfer* version 6.0, a free open-source software used to quantify brain anatomy. *FreeSurfer* encompasses multiple brain atlases to produce brain segmentation. We selected the Desikan-Killiany-Tourville atlas^42^ (DKT, aparc.DKT.stats file) for the cortical segmentation. The MRI data were processed using the “recon-all -all” command in *FreeSurfer* with the fully automated directive parameters (no manual intervention or expert flag options) on the CBRAIN^43^ platform to extract thicknesses for all defined atlas regions.

The processing techniques used by FreeSurfer^18^ to generate the cortical models consisted mainly of motion correction, removal of non-brain tissues using watershed/surface deformation procedure, automated Talairach transformation, intensity normalization, tessellation of the gray matter white matter boundary, automated topology correction, and surface deformation following intensity gradients to accurately place the tissue boundaries between gray/white matter and gray/cerebrospinal fluid. Additional processes are available following the cortical model : surface inflation, registration to a spherical atlas and parcellation of the cerebral cortex into units. These allow for a better representation of cortical thickness by using the intensity values and the whole information from the volume segmentation. Thickness is calculated using the closest distance from the gray/white boundary to the gray/CSF boundary at each vertex on the tessellated surface^33^.

For this project, we used the average regional cortical thickness per region as a proxy of cortical neuronal degeneration, as validated in the work of Cardinal and Belathur^44,45^. This average was turned into a Z-score using the NOMIS tool, which normalizes the data based on a two-step regression model taking into account image quality and head size (eTIV), and sex. Since age is an important factor in AD it was not used here to normalize data. Hence, for each participant, expected measure values were generated in the form of Z-scores that demonstrate deviations from the normative NOMIS values.

### Data analysis

#### Low dimensional reduction

According to the DKT atlas, the cortex was divided in 31 regions for each of the left and right hemispheres, and hence each data point was represented by a 62-dimensional vector. To facilitate predictive modeling, dimensional reduction of the data with UMAP was an important step. UMAP is a nonlinear algorithm based on Riemannian geometry and algebraic topology used for dimension reduction of high-dimensional datasets. To accomplish this, UMAP first computes the topology of the manifold as a *k*-nearest neighbor graph. The graph is weighted by the probability that two points are connected based on their distance computed with the Riemannian metric. The further two points are from each other, the least probable they are to be connected. For a given node, the number of nearest neighbors with which distances can be calculated are determined by the hyperparameter `n_neighbors`, denoted by *k* in the mathematical and computer science literature^34,35^. The other hyperparameter, `min_dist`, describes the minimum distance between points in the reduce space. When, UMAP tries to project the data from a high Riemannian space onto a lower Euclidean space, it does so by respecting the minimal possible distance between points in that space. Finally, the topological graph is generated for the low-dimension space, and by optimizing the cross-entropy between the lower and higher dimension topology, the algorithm finds the projection that represents the most the data topology^46^(further details are included in McInnes^34^). Has previously mentioned, the two hyperparameters give the algorithm generalizability while allowing us two find the embedding that fits our need. In our first experiment, we generated a low-dimensional manifold using UMAP by reducing the 62-dimensional cortical thickness data from NOMIS to *m*-dimensions (with its axes being UMAP0, UMAP1, …, UMAP*m*− 1), where *m «* 62, using `min_dist` = 0 and different `n_neighbors` values of 2 to 100 (Figure 3, Figure 4). We initially verified if *m* = 2 or *m* = 3 was the better target dimension for our problem. To do so, the explained variance of each dimension was calculated to verify if the third dimension was useful. We then used the trained model to transform the entire ADNI data in the new *m*-D space.

**Figure 2.**
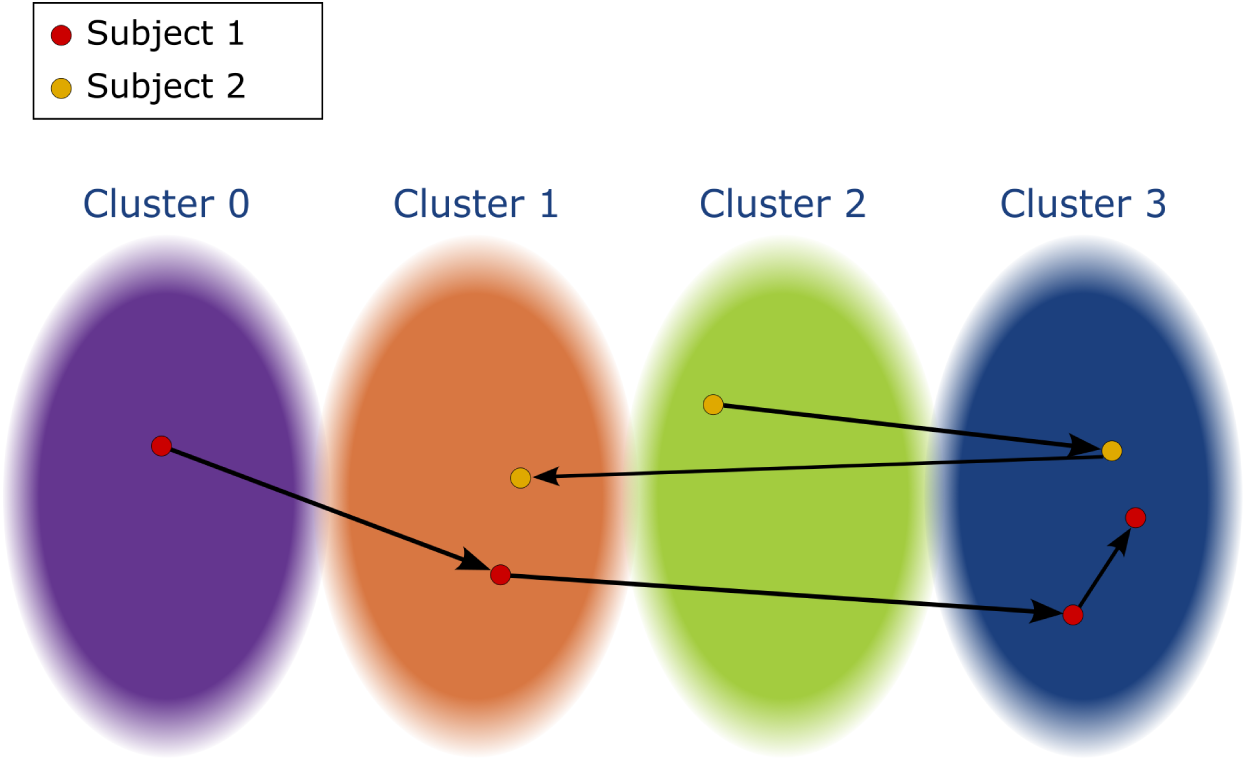
Defining trajectories. An arbitrary embedding with four clusters. The arrows represent the direction in time for each subject. Given an embedding with clusters we can define the longitudinal trajectory of subjects as follows. In this example, subject 1 would have a full trajectory of [0-1-3-3] while subject 2 [2-3-1]. The trajectory of length 1 would be [3] for subject 1 and [1] for subject 2 and the trajectory of length 3 would be [1-3-3] for subject 1 and [2-3-1] again for subject 2. Subject 2 wouldn’t have a trajectory of length 4 and subject 1’s would be equivalent to its full trajectory.

**Figure 3.**
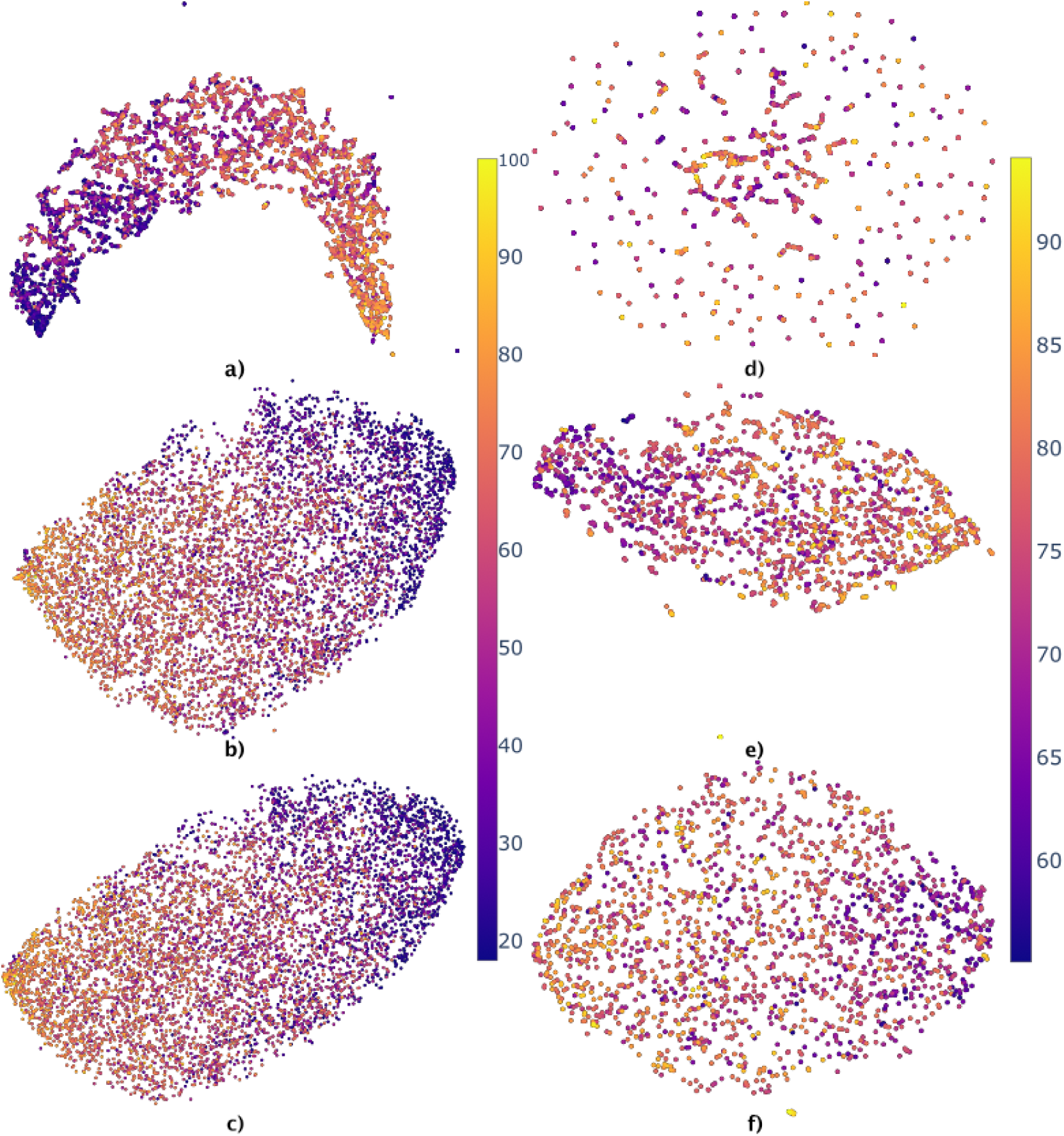
UMAP embeddings of the NOMIS (left figures) database and ADNI database (right figures) using three different values of `n_neighbors`. The age (years) of the participants is color coded, from 18 to 100 for NOMIS and 55 to 97 for ADNI. The parameter `min_dist` = 0 for all embeddings. Figure 3a) and **d)** has `n_neighbors` = 3, Figure 3b) and **e)** has `n_neighbors` = 20 and Figure 3c) and **f)** has `n_neighbors` = 100. Note that axes have been scaled to fit every point in the frame.

**Figure 4.**
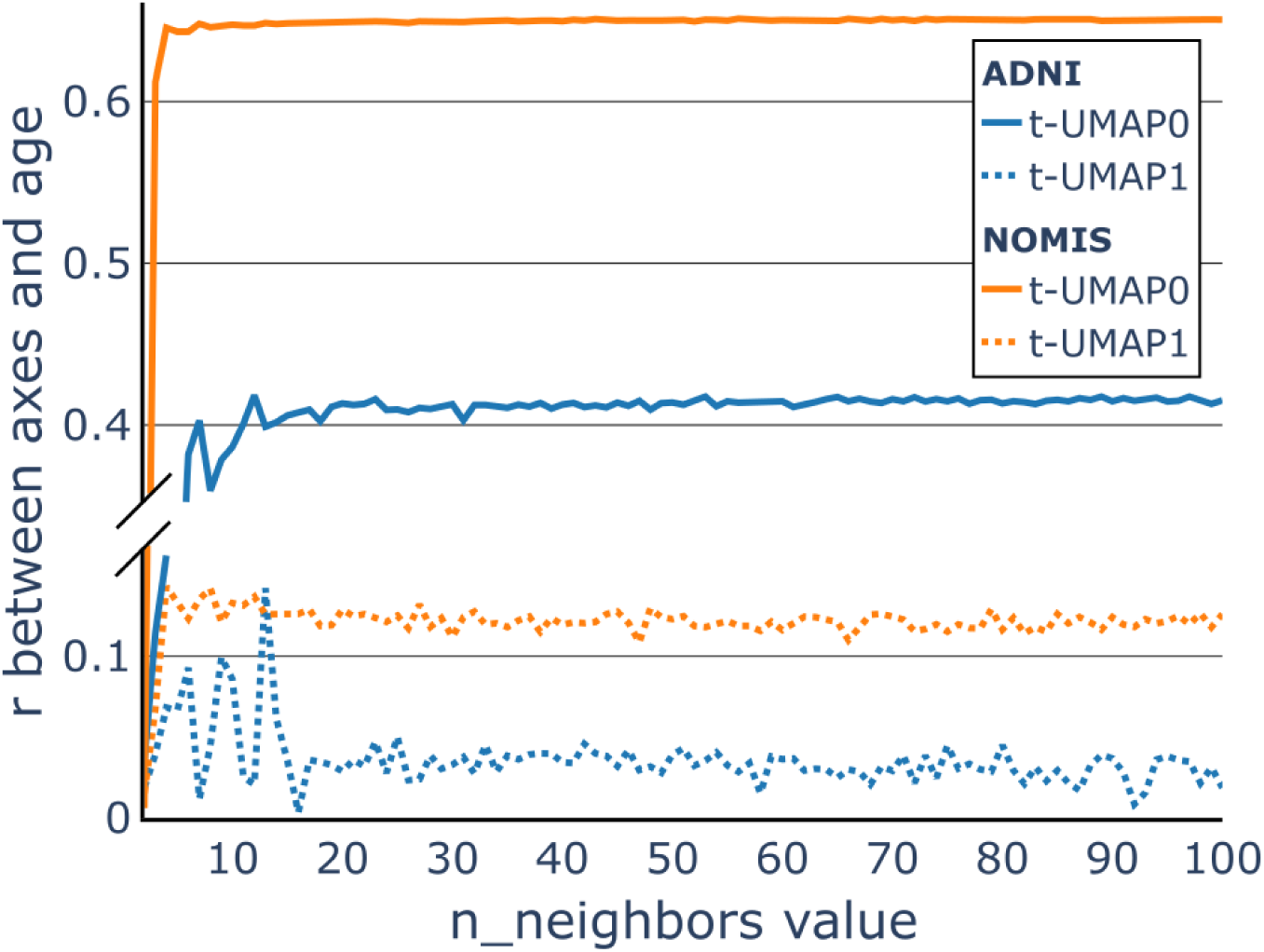
The choice of hyperparameter. The Pearson correlation (*r*) between the values on the t-UMAP0 axis (full line), the values on the t-UMAP1 axis (dashed line) and the age of corresponding participants in the NOMIS dataset (orange) and ADNI dataset (blue). The correlations are plotted against the value of the hyperparameter `n_neighbors` used to generate the t-UMAP embedding. This allows us to find the value where the correlation is greater.

In our second experiment, we created a low dimension embedding model using the ADNI database only. For this model, we randomly selected 40% of the subjects in the ADNI dataset to generate the UMAP embedding, in which we projected data from the remaining participants, performed clustering, and calculated the probability of AD associated with trajectories. For both models, no feature selection was made on the data so that UMAP has full information to maximize the distance between MCI-MCI and MCI-AD and see if relevant cortical regions can be extracted.

We hypothesized that the first model would encompass the characteristics of normal aging which could distinguish AD and MCI patients, but the second model could directly represent the morphological decline related to dementia and thus be more predictive of cognitive outcome. In the rest of the paper, we will use *NOMIS embedding* to refer to the embedding trained on NOMIS and *ADNI embedding* to refer to UMAP models trained on ADNI.

#### Transformation for axis analysis

In this application of the nonlinear UMAP method, it is difficult to interpret axes in terms of the features (thickness of cortical regions). Remark that algorithms such as PCA provide a linear combination of features to generate axes, while with UMAP these relations come from correlations. Moreover, the UMAP embedding doesn’t impose an orientation on the data and thus, any rotation is equally valid. Therefore, after having applied the UMAP algorithm, we used a linear transformation over the UMAP embedding to rotate the data such that its principal axes could explain most of the variance in the embedding. As described by the principal component analysis, this rotation is given by the singular value decomposition of the embedding^47^. This method allowed us to determine which features correlated the most with UMAP embedding axes. These transformed axes are referred to as t-UMAP. Because aging is a strong confounder in the study of dementia, we tested whether the algorithm implicitly embedded age in both NOMIS and ADNI embedding representations. To test this, we used Pearson’s correlation between age and t-UMAP vectors. We further tested if there were correlations between the axes and the thickness of the different brain regions.

#### Clustering

In either embedding experiment, the low-dimensional t-UMAP space was then parcellated into data clusters, each containing the same number of data points, in order to achieve similar statistical power in our trajectory analyses. The clustering was done on the t-UMAP coordinates only. To this end, we adapted *scikit-learn*’s *k*-means algorithm^48^ to our need of segmenting the data in equal groups.

A number of clusters (*C*) can be specified by the user and the number of data points in each cluster (*n*) is determined from the overall size of the data set. The clusters’ centroids were initialized using *scikit-learn*’s *k*-means++ algorithm^49^. A distance matrix between each pair of points and centroids was then computed, and a probability matrix calculated. The entries of the latter matrix describe the probability that a given data point (row) belongs to a given cluster (column). These probabilities are based on the distances between data points and the centroids according to the formula: **W***_i j_* = 1 − **D***_i j_/***V***_i_* where **W** is the probability matrix, **D** is the distance matrix and **V** is a column vector where each element is the maximum value of the corresponding row in **D**. The matrix entry has a value of 0 if the corresponding centroid is the furthest from the data point.

For each cluster, the *n* likeliest points were then assigned to it. If the number of data points was not divisible by *C*, the unassigned data points were assigned to their likeliest cluster.

An iterative process followed, in which, at each iteration, centroids’ position and their distances from every point were calculated; probabilities for a point to switch cluster were ordered; and assignments performed. The process was run until the inertia (within-cluster sum-of-squares of the distances between points and their closest centroid) could not be minimized further, or a maximum number of iterations was reached^48^.

### Probabilistic Trajectory

In the *m*-dimensional space computed from the embedding data, we projected the remaining ADNI data points and segmented them into identical-size clusters, after which we turned our attention to the problem of trajectory prediction.

We first defined as a trajectory the ordered sequence of clusters through which a participant’s data belonged in its longitudinal course. Figure 2 shows a simplified representation of an embedding with four clusters. As shown in the figure, we defined a subject’s trajectories from their latest data point, meaning a trajectory of length 3 is composed of the 3 oldest datum for a subject. To characterize trajectories at the group level, in every possible trajectory, we computed the probability that an individual in this trajectory developed AD. First, given an embedding with *C* clusters, the total number of trajectories *T*, for every possible trajectory of length one to *L*, is given by 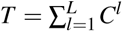. Each set of *T* trajectories is initialized with a null probability. The probabilistic atlas was then generated, in which every entry was composed of a trajectory and its associated AD progression probability. Then, for every participant, we pieced together cluster trajectories for the last, two last and three last data points, thus creating trajectories of length one, two and three steps for the participants. Depending on the final diagnosis at the last step (i.e. whether the participant remained stable or progressed to AD), the probability of this trajectory with respect to AD was updated consequently. Once the whole dataset was processed, we had an atlas tying the probability of developing AD to each possible trajectory of lengths 1 to 3.

A disadvantage of this approach is that empty trajectories (paths not taken by any participant) were evaluated as paths with a probability of zero, even though no data were available. These trajectories cannot be tossed aside since future participants might follow such trajectories, and a statistic will be required to “predict” diagnosis. We then arbitrarily chose for these trajectories to remain at a value of zero.

Given that ADNI has 949 MCI individuals at baseline, *C^l^* must be *«* 949 so that trajectories of length *l* are reasonably populated. Because of this limitation, we empirically limited the length of a trajectory to the last three data points (i.e., three years before diagnostic of AD) and the maximum number of clusters to 7, which gave us a maximum of 343 unique trajectories. Since the subjects are not uniformly distributed between every trajectory, as shown in Supplementary Figure S5 and S6, we could afford *C^l^* to be *≈* 1*/*3 of the number of subjects. Indeed a lot of trajectories will never be used and can thus be discarded. Supplementary Figure S5 shows that *C* = 7 and *l* = 3 gives 108 used trajectories. Given larger datasets, these limitations could be lifted.

### Statistical analysis

We performed a sensitivity analysis using 10-fold stratified cross-validation as follows. First, we separate the ADNI dataset into three groups : the test set (10% of total MCI-MCI and 10% of total MCI-AD subjects), the space set (40% of total MCI-MCI and 40% of total MCI-AD subjects) and the cross-validation set. The latter is then separated using 10-fold stratified cross-validation.

For the NOMIS embedding, only the test and cross-validation set were used. These groups were projected in the transformed UMAP space, created by the NOMIS dataset. One of the fold was chosen as the validation set and the rest were used as the training set. The cluster segmentation algorithm was then applied on the training group, and the probabilistic atlas is calculated as defined above. We then extracted the final results using the validation group. For each participant, we calculated the probability of progressing to AD given their last 1, 2 and 3 data points. We then set an arbitrary threshold to binarize predictions (e.g., every prediction with a probability higher than a threshold of 0.5 would be qualified as an AD subject). We computed a receiver operating characteristic (ROC) curve for each trajectory length (1, 2, or 3) by varying the value of this threshold from 0 to 1, calculating the true positive rate (*sensitivity*), false positive rate (1 *− specificity*) and area under the curve (AUC). The process is done on different values of `n_clusters`. The best model was chosen from this and was then applied to the test set to calculate the actual accuracy of the model.

As for the model generated from ADNI embedding, we used the exact same test, space, and cross-validation sets. In this case, the space set was used to generate the ADNI embedding. Following this embedding, the test and cross-validation set were reduced and the latter separated using 10-fold stratified method. From that point, the same cross-validation process was applied. Finally, the test set was clustered using the trained model and a sensitivity analysis performed.

## Results

### UMAP transformation

Using UMAP on the standardized NOMIS and ADNI data, we explored different embedding possibilities as a function of `n_neighbors` values (Figure 3). For the NOMIS dataset, each embedding encompasses a strong correlation between the first (UMAP0) axis and age, as shown by the color gradient. However, for high `n_neighbors` values, changes in the embeddings were less prominent. For instance, Figure 4 shows that the correlation between t-UMAP0 and age varies only slightly after `n_neighbors` = 5. On the other hand, for the ADNI embedding, changes were much more prominent. By changing `n_neighbors`, points were more and more uniformly distributed along the space. Moreover, the participants’ longitudinal scans were closely grouped up in embeddings with lower `n_neighbors` (Figure 3**d)** and **e)**). This result reflects the choice of `min_dist`=0 since it makes feature vectors resembling themselves as close as possible in the UMAP space. On the other hand, age was not well characterized in these models with `n_neighbors`=3 showing no signs of age-related gradient and `n_neighbors` >= 20 only displaying clear differences between the extremities.

To select the embedding for our trajectory analysis, we computed the correlation between t-UMAP0 (full line) and t-UMAP1 (dashed line) with the age of subjects from NOMIS (orange) and ADNI (blue) when varying the parameter `n_neighbors` (Figure 4). For the NOMIS model, t-UMAP0 at low values of `n_neighbors`, the correlation steeply increases before quickly converging towards an asymptote. The correlation between age and t-UMAP1 also seems to converge to an asymptotic value albeit in a more noisy fashion. Hence, to choose the optimal `n_neighbors` value we wanted to maximize correlation between t-UMAP0 and age, while minimizing the one between t-UMAP1 and age. Optimal correlations were contained between `n_neighbors`=20 and `n_neighbors`=47, meaning that differences were most likely due to random noise. Therefore, to preserve computer resources and the local structure of the underlying manifold, we chose `n_neighbors`=20.

For the ADNI model, the models were not strongly correlated with age with a maximum *r* just over 0.4 for t-UMAP0 and around 0.05 for t-UMAP1. Even though these models do not seem to be related to biological age, we chose `n_neighbors`=16 since the correlation with t-UMAP1 is at its minimum and starting to plateau for t-UMAP0.

We also tested higher dimensional embeddings. Using a 3-dimensional embedding with `n_neighbors`=20, we computed the explained variance of each dimension to quantify the usefulness of each dimension. The first dimension explained 95.91%, the second 2.93% and the third 1.16% of the variance. We felt therefore justified in working with only the first two dimensions.

### Reorientation of UMAP embedding along its principal axes and correlations

In our first experiment, once the embedding’s axes were aligned with the principal axes, we noticed that participants’ ages correlated with the transformed main UMAP axis (t-UMAP0; *r* = 0.65) (Figure 5). It has to be noted that chronological age was not used as an input in the UMAP transformation and this is therefore an emergent property of the embedding. There were six brain regions that were heavily negatively correlated with t-UMAP0 : superior frontal gyrus (*r* = −0.88), supramarginal gyrus (*r* = −0.85), inferior parietal gyrus (*r* = −0.83), rostral middle frontal gyrus (*r* = −0.83), caudal middle frontal gyrus (*r* = −0.82) and superior temporal gyrus (*r* = −0.81). As for t-UMAP1, the peri-calcarine (*r* = 0.55), the cuneus (*r* = 0.48) and the lingual gyrus (*r* = 0.47) were the three regions with the highest correlation. One notices in Figure 5 how the older ADNI participants (age at entry > 65 y.o.) preferentially concentrate along the positive t-UMAP0 direction, congruent with its correlation with age in the NOMIS dataset.

**Figure 5.**
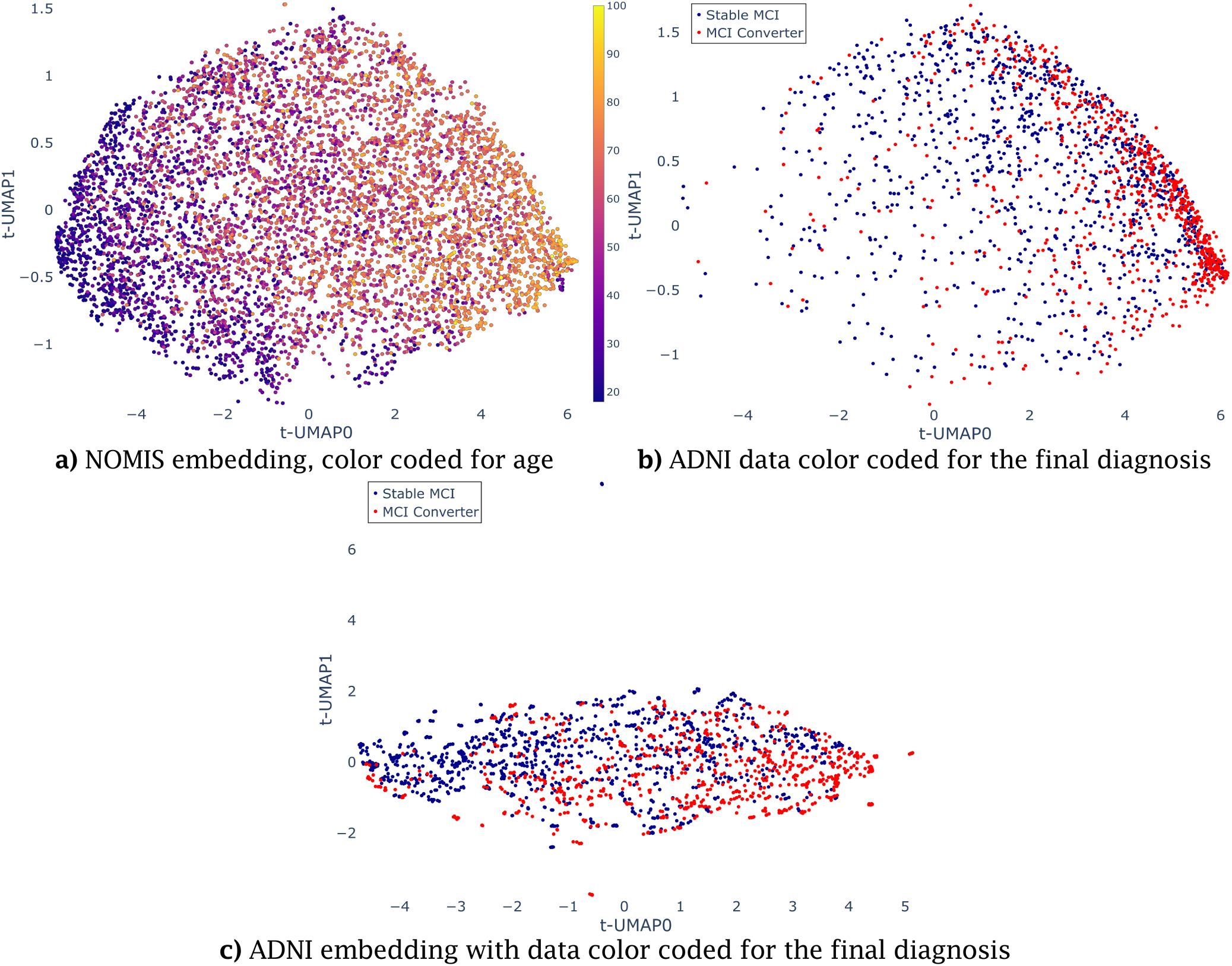
Transformed UMAP embedding and fitted ADNI data. A linear transformation is applied on the UMAP embedding with `n_neighbors` = 20 and the ADNI embedding with `n_neighbors` = 16 to align the principal axes with UMAP0 and UMAP1. In **a)**, the NOMIS database (embedding data) with participants’ chronological age (years) at scan color-coded using the color scale displayed on the right. In **b)**, participants from the ADNI database were projected in the NOMIS-defined UMAP embedding. Subject’s final diagnosis was used to color the points on **b)**. One notices how the older ADNI participants (age at entry > 65 y.o.) preferentially concentrate along the positive t-UMAP0 direction, congruent with its correlation with age in the NOMIS dataset. In **c)**, subjects from ADNI are reduce using UMAP and color coded by the final diagnosis.

For the ADNI embedding, Figure 5**c)** shows different patterns. First, the AD population is once again concentrated to the right, but in a less aggregated manner, implying that although the AD population moves towards the right of the figure, there are no clear attraction points, when compared with the NOMIS embedding. Again, the MCI-MCI are mostly uniformly distributed over the space. The data does not form well-defined clusters, but interestingly they tend to group up in small aggregates of less than 10 points that can look like a single point on Figure 5**c)** (for example the singular point around (0,7) that is in reality 6 points). The data in these aggregates usually come from the same patient. Interestingly, the correlations for the ADNI embedding are less significant. Most correlations were smaller than previously observed with the four most correlated brain regions being supramarginal gyrus (*r* = −0.81), superior temporal gyrus (*r* = −0.76), superior frontal gyrus (*r* = −0.75) and middle temporal gyrus (*r* = −0.74). The age is no longer correlated (*r* = 0.4). For t-UMAP1, only the entorhinal cortex stands out (*r* = 0.54), with the other regions being mostly uncorrelated (0.3 *> r > −*0.3) and the same is true for the age (*r* = 0.003).

For further information, consult Supplementary Tables S1 to S4 for the full extent of the correlations between cortical regions and t-UMAP0 and t-UMAP1.

### Trajectories

In our first experiment, to get the trajectory of each individual, we first separated the ADNI dataset into the training data, projected it in the NOMIS t-UMAP space, and separated this embedding into clusters. In Figures 6 and 7, we used the same_size cluster algorithm defined in the section *Clustering* with `n_clusters`=4.

**Figure 6.**
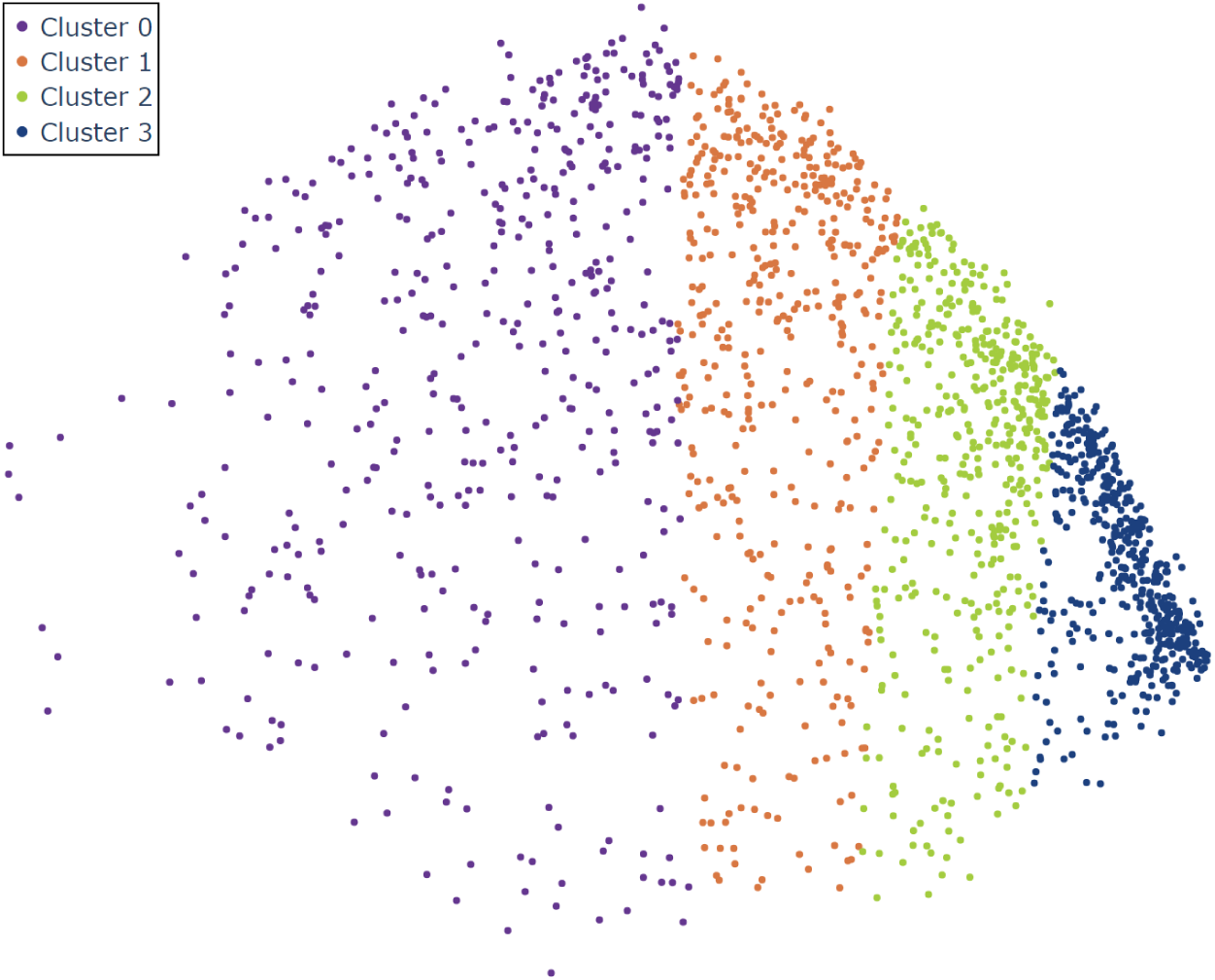
Clustering of the ADNI embedding with same size *k*-means. Dimensional reduction of the ADNI database using a linearly transformed UMAP model trained on the NOMIS database using `n_neighbors` = 20 and `min_dist` = 0. The four colors represent the clusters made from the positional values, using a variation of the *k*-means algorithm with `n_clusters` = 4. For simplicity’s sake, the axes are not displayed since they are the same as Figure 5.

**Figure 7.**
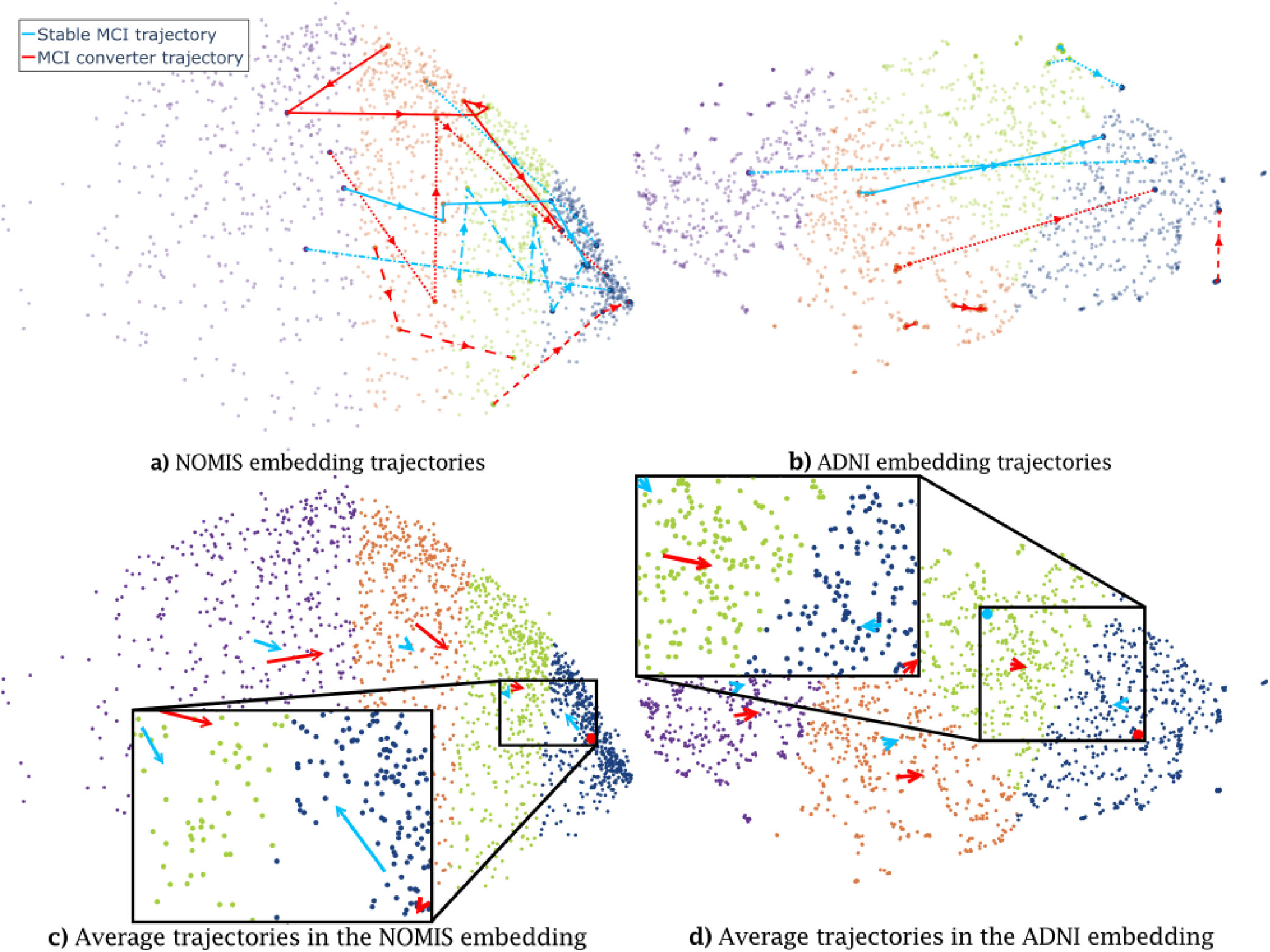
Individual and average trajectories for MCI-MCI and MCI-AD subjects for the NOMIS (left) and ADNI (right) embeddings. On top, the individual trajectory of four MCI-MCI and four MCI-AD patients in the NOMIS embedding (Figure 7a) and the ADNI embedding (Figure 7b). Different dashed lines are used to differentiate between different subjects, within the same diagnosis, and the arrows point in the direction of time. The same patients are shown in both figures. Figure 7c) and d) shows the average trajectories for both types of trajectories. Some vectors are represented by a point because the trajectory is too short to be represented by an arrow. Figure 7b and d have had their t-UMAP1 axis cropped to exclude extreme values and visualize trajectories seamlessly.

For illustrative purpose, we show in Figure 7 how different participants move through the embedding with time (a “trajectory”). ADNI participants tend to move in a positive direction along t-UMAP0, i.e. from clusters on the left (clusters 0 and 1) to those on the right (clusters 2 and 3) as they age. Some subjects have a direct trajectory to the right-most cluster (solid blue line in Figure 7a), while others have a more convoluted path (dotted red line in Figure 7a). On the other hand, some participants move from right to left (not displayed on 7, see 8), in the negative t-UMAP0 direction.

For the ADNI embedding(Figure 7b), trajectories are less chaotic and most move in a straight direction. Some trajectories, like the full red line, still loop around and some others have less linear paths, but they are much smaller in scale as compared to the linear trajectories. Moreover, some trajectories are much shorter, which is reflected by the fact that the same patient’s scan are often in the same neighborhood.

Figure 7c demonstrates the trends for the average MCI-MCI and MCI-AD groups. Notably, MCI-AD subjects whose trajectory initiates in cluster 3 tend to remain in this cluster, while MCI-MCIs move back towards cluster 2. Also, MCI participants whose trajectory initiates in clusters 1 and 2 display shorter movement towards the right for MCI-MCIs and a greater movement in this direction for AD. Thus, there seems to be an attraction towards a center of mass in cluster 3 for ADs, and in between clusters 1 and 2 for MCI-MCIs.

For Figure 7d, the average trajectories are overall significantly shorter than those from the NOMIS embedding. We see that MCI-MCI average trajectories are always shorter than MCI-AD’s (excluding the final cluster) and AD trajectories do not exert significant movement in cluster 0 and 1, but all averages are still moving on the t-UMAP0 axis in a positive direction. This can again be explained by the fact that many patient’s scans are grouped closely together, making the averages shorter also. The last cluster seems to again attract AD subjects, but MCI-MCI subjects do not move significantly away from it for this embedding.

Figure 8 displays the probabilistic atlas in its entirety. Using the embedding and clusters from Figure 6, we computed the proportion of individuals having converted to AD in each possible trajectory of length 1, 2 and 3, using participants’ last *l* data points. This allows us to make some further observations on the trajectories. First of all, in Figure 8a there seems to be a relationship between the last cluster being visited and prognosis. For instance, trajectories 0 ; 0-0 ; 0-0-0 have a very low probability of developing AD (*<* 18%), while trajectories 3 ; 3-3 ; 3-3-3 have a probability between 62% and 77% to be converting MCIs, with intermediate clusters displaying a progressive relation. Secondly, out of the 56 trajectories with at least one subject, 30 were monotonically increasing or stable (i.e. moving from lower to higher cluster ID or staying in the same cluster). These seem to follow the previous phenomena related to aging and eventual conversion to AD. The second half of trajectories (n = 26) were more random. However, the population distribution for these trajectories have a median of three subjects; while the five trajectories with 10 or more subjects all had an associated conversion probability of under 42%. Thus, they seem more representative of either noisy data or morphological variability without association to conversion. Moreover, trajectories that have a point in cluster 3 without finishing in it have all probabilities ≤ 50% and are more populated than most of the abnormal trajectories ; with a median at 4, a mean at 6.6. This shows that these trajectories are more often associated with MCI even though this cluster is by itself more associated with the AD. Finally, there are a number of outlier quadrants with either a probability of zero or one, due to a limited sample size (1 to 3 subjects).Figure 8b shows a similar pattern. Indeed, we see again that patients in the last cluster have a greater probability of developing AD with this probability augmenting the longer the stay (68% to 74%) while patients in the first cluster have the lowest probability associated (*<* 17%). On the other hand, the in-between clusters behave differently. Subjects ending in cluster 1 have a higher probability (38%-41%) than what was observed in the NOMIS embedding and cluster 2 patients have a lower probability (28%-31%) than cluster 1.

**Figure 8.**
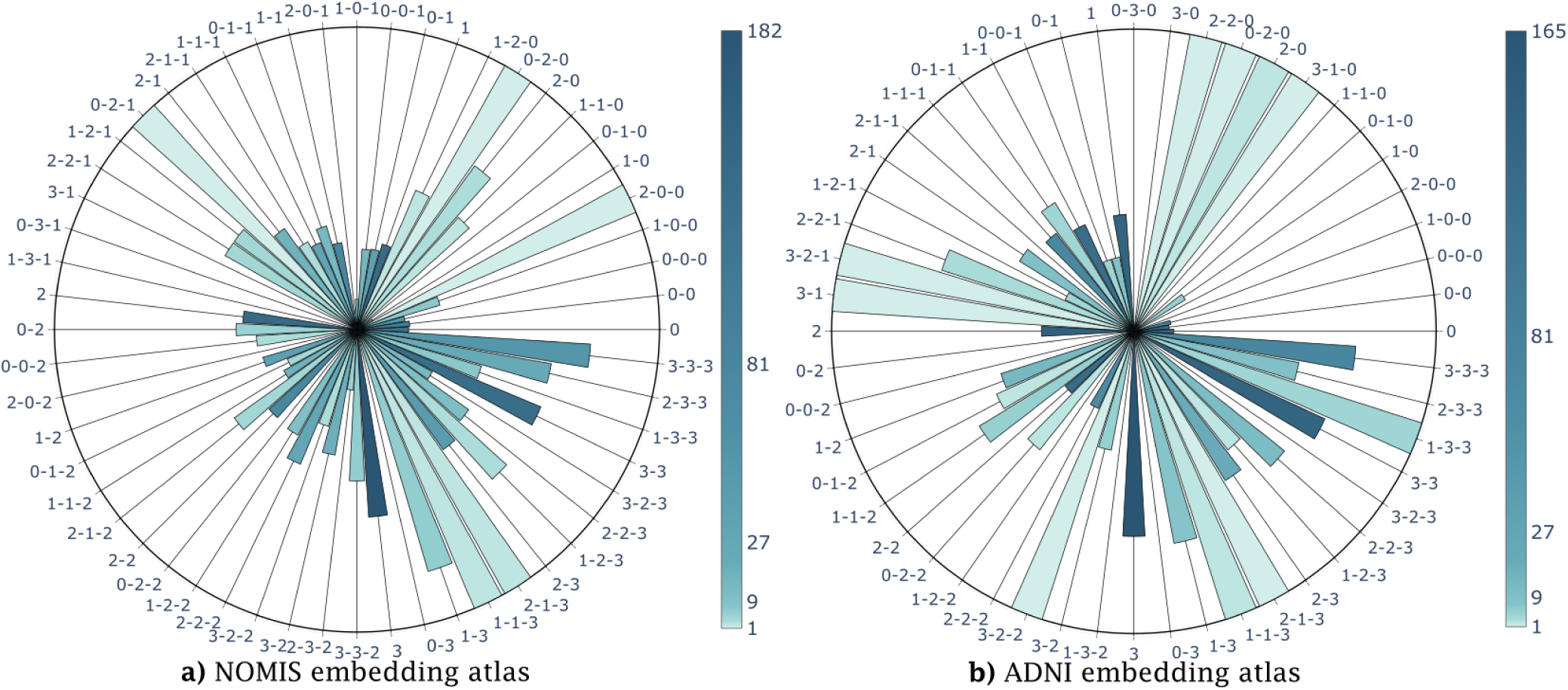
Probabilistic atlas. The distribution of the proportion of AD subjects in each trajectory of length 1 to 3, color coded for the population in each trajectory. For the sake of visibility, unpopulated trajectories are not displayed. These charts are obtained by counting the number of MCI-MCI and MCI-AD participants in each trajectories. The full bars represent trajectories containing only MCI-AD participants while empty bars have only MCI-MCI participants. Figure 8a) is the atlas for the NOMIS embedding and **b)** the atlas for the ADNI embedding.

### ROC curves

We computed the AUC of our ROC curves as an indication of the efficiency of our probabilistic atlas at classifying the test group. We first randomly divided ADNI into a test set (10% of every MCI-MCI and MCI-AD), space set (40%) and a set for the validation process (50%). The latter was then separated into a training group, containing 45% of every MCI-MCI and 45% of every MCI-AD participant, which we used to compute the probabilistic atlas for trajectories of length 1, 2 and 3, with the previously mentioned dimensional reduction method and clustering method. 5% of data left was used to validate the accuracy of our method. Varying the threshold probability value at which the participant was deemed to convert allowed us to generate ROC curves shown on Figure 9.

**Figure 9.**
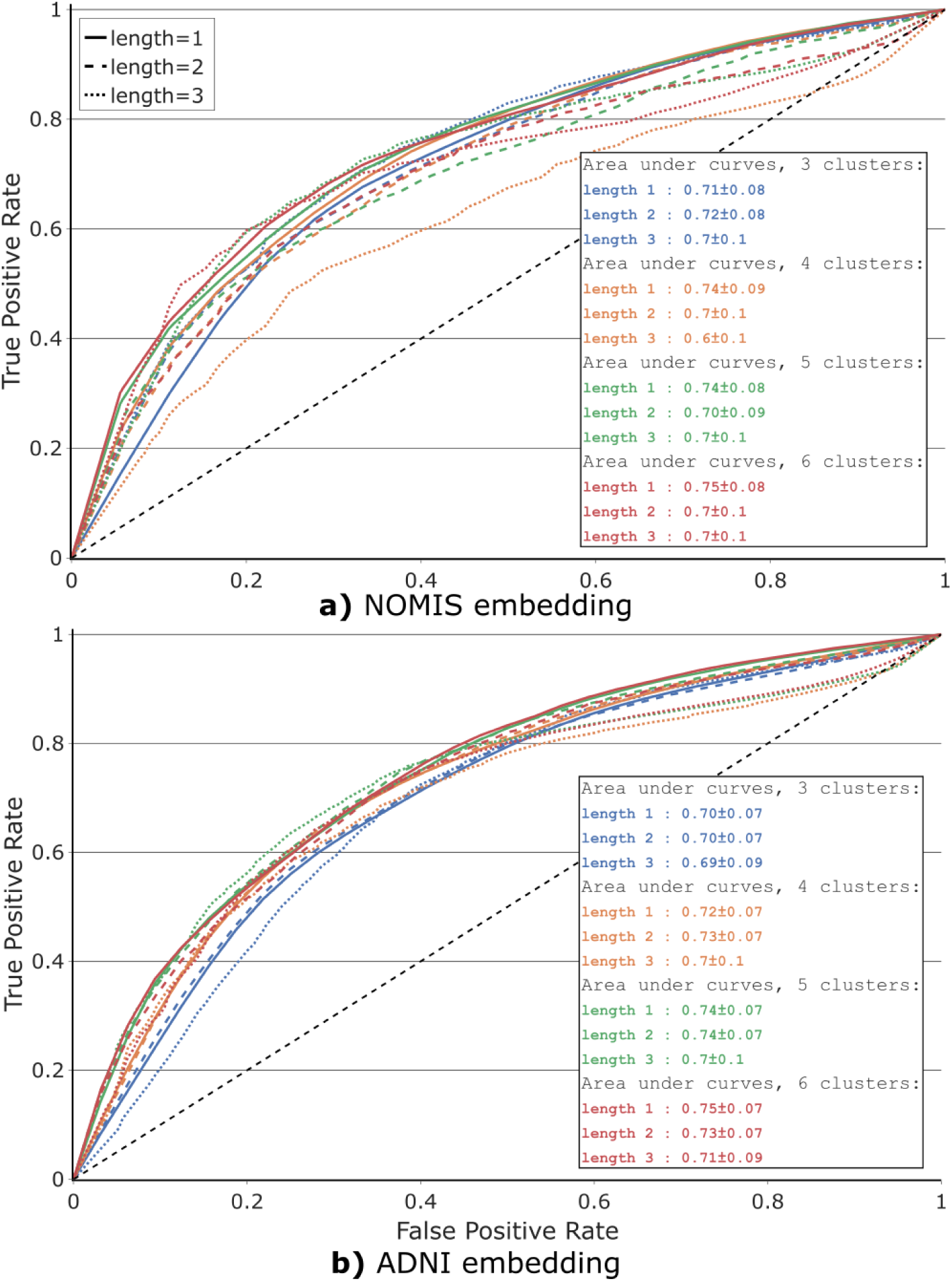
ROC curves of 4 different clustering of the transformed UMAP embeddings (with 3, 4, 5, and 6 clusters). The ROC curves are generated from the classification between MCI-MCI and MCI-AD trajectories probabilities from the probabilistic atlas (Figure 8). For each clustering, we verify the effect of trajectory length, the last *l* longitudinal time points of a participant; see Figure 2. Figure **a)** shows the ROC curves obtained using the NOMIS embedding, while Figure **b)** is made using the ADNI embedding. Both figures use the same cross-validation set (subset of ADNI). For each clustering, predictions are made using trajectory length of 1, 2, and 3 last time points. The dashed black line represents the diagonal with an area under curve (AUC) of 0.5 for reference. The AUC of each ROC curve is displayed in the table at the bottom right of the figures. This value is obtained by averaging the AUC from the 10-fold resamplings of the train/validation subsets and the value following the *±* is the standard deviation of this distribution. Moreover, the ROC curves displayed are obtained by averaging the ROC curves from the same 10-fold resamplings.

The training and validation sets were resampled using 10-fold cross-validation which is repeated 10 times with different random states. The average and standard deviations for this AUC distribution were calculated and are displayed as an insert in Figure 9. Accuracy grows as a function of `n_clusters` for a given length, with the most accurate models at `n_clusters`=6 for the NOMIS embedding and ADNI embedding, but high variance on length 3 makes this relation difficult to confirm. The algorithm was also tested on 7 and 8 clusters, but the AUC plateaued for those. Alongside higher standard deviations caused by the increasing number of empty trajectories (Supplementary Figure S5 and S6), these models were excluded for simplicity.

Finally, the test set was applied on the NOMIS and ADNI model that yielded the best AUC during validation. From those we got a final accuracy of 0.72 for the NOMIS embedding with 6 clusters and trajectory length of 3 and 0.80 for the ADNI embedding with 4 clusters and trajectory length of 3.

## Discussion

Our goal in the present work was to investigate low-dimensional representations of aging trajectories in order to identify differences between trajectories related to stable cognition and decline due to probable AD. We wanted to develop a method that has a good predictive accuracy, but at the same time, remains interpretable. To do so, we first extracted a vector of cortical thicknesses of different brain regions using FreeSurfer. We then further reduced the dimensionality of the data using the nonlinear method UMAP and linearly transformed the UMAP embedding while maximizing the correlation of the main axis with age. We then investigated the different aging trajectories in this space and found significant differences between MCI-MCI and MCI-AD trajectories. We estimated that our novel approach could yield a good predictive accuracy, with a maximum AUC of about 0.72 on the NOMIS embedding and 0.8 on ADNI embedding. We now discuss the novelty of our approach, its limitations and possible future research directions.

### UMAP as a nonlinear embedding technique

UMAP was chosen over PCA since the former provided a more compact and homogeneous embedding of the data, with a lesser standard deviation in both the *x* and *y* axes, which is advantageous when dividing the space into clusters of similar size. The nonlinearity of UMAP is advantageous as it represents more accurately cortical thickness atrophy happening with age, while preserving both local and global features of the original high-dimensional dataset. The optimization of UMAP parameters required us to fix `min_dist` to 0. This means that two MRIs yielding the same image will occupy the same position in the t-UMAP space. This is useful since it will induce a positive distance between two points only if they are different, and thus a participant’s trajectory is not artificially modified. Furthermore, two similar points are more likely to be in the same cluster. Then, for the parameter `n_neighbors`, Figure 3 showed the different embeddings given a value of the parameter. As explained in the UMAP paper^34^, lower values make the embedding locally connected while higher values make it globally connected, meaning that global structures in the data are represented over local ones. The figure represents this by showing the softening of internal structures for high `n_neighbors` values. Indeed, for the lowest value of `n_neighbors`, the embedding from NOMIS takes a very different form. With low values, local resemblance between members of the NOMIS database are put forth while the more `n_neighbors` is high and the more the embedding starts looking alike : points are uniformly distributed inside an ellipsoid. This is even more apparent for the ADNI embeddings where, on low values, the data is regrouped in tightly packed clusters to the point of resembling to a single data point in some case. Increasing `n_neighbors` also causes this embedding to take an elliptical form, while conserving a level of local connectivity. This connectivity comes most likely from the fact that ADNI is a longitudinal database as opposed to NOMIS. Since the data is standardized, a scan should be located in the vicinity of its subject’s other scans in the UMAP space, unless a significant change happened in the morphometry. In the embedding, this is reflected by a locally connected plot where each subject’s scans are often regrouped together in the space, even with high `n_neighbors`. For the current paper, we prioritize global structures since embeddings with lower `n_neighbors` are less suitable for the same reason PCA was not. Moreover, we chose the correlation between t-UMAP0 and subject’s age as a metric of how much the embedding encompasses the aging process. Figure 4 shows that this value is optimized from `n_neighbors` = 20 for the NOMIS embedding and `n_neighbors` = 16 for the ADNI embedding. In light of this, we chose the embedding with `n_neighbors` at 20 and 16 since it offered a good compromise between local and global structures and the correlation with age is optimized for all axes (Figure 4). Finally, before settling on a two-dimensional representative space, we verified that a higher-order embedding would not better fit the data. The explained variance of the third dimension (1.16%) showed that the first two dimensions were sufficient to represent our datasets.

In insight, the correlation with age might not be the most accurate metric for choosing `n_neighbors`. Although it makes sense for the point of view of having an embedding representing aging, the fact that the ADNI embedding has very low correlations with age and yet manages to obtain better predictive accuracy shows that this metric doesn’t paint a full picture of the embedding’s predictive power. Future studies should either find a relevant measure to evaluate the embedding, or if computational power is not a problem, include `n_neighbors` as a parameter of the cross-validation process.

### Age dependency

Applying a linear transformation on the embedding in order to align the principal axes with the UMAP0 and UMAP1 axes yielded a new space where the t-UMAP0 axis correlated the most with the age. Indeed, the age gradient on Figure 5 seems to progress parallel to the axis with younger individuals are located on the left of the figure while older ones are on the right. The t-UMAP0 axis can be interpreted as a participant’s “brain age”; the fact that some younger participants had a strong “positive” brain age is indicative of individual variability. Note however that *FreeSurfer* seems to introduce significant noise in morphometric measurements, which causes highly variable trajectories for some individuals.

In this paper, we specifically chose to use a dataset of healthy individuals with a wide age range to train the UMAP embedding. This was done with the idea that the embedding would encompass the normal aging process which would make it easier to differentiate AD trajectories down the line. In a way, it is what we were able to do since the embedding correlates with the brain age of subjects. Interestingly enough, the data is relatively uniformly distributed in the embedding and no clearly defined clusters are present. This shows again that UMAP wasn’t able to find defined characteristics in the data and so every subject is represented in a spectrum of the aging process. Moreover, we chose to use no prior to select the features to be reduced. This again goes with the idea we wanted the embedding to represent the aging process of the brain. For the specific purpose of predicting decline, we could have used *Dickerson* et *al.*’s cortical AD signature^7^ to select specific AD related regions, which would have helped increase the sensitivity of our model and hence its predictive power. On that note, the inferior parietal and superior frontal gyrus are both heavily correlated with the t-UMAP0 axis and have been identified as regions within the AD signature^7^. On the other hand, the ADNI embedding doesn’t express similar correlations. The correlations are all lower as compared to the NOMIS embedding (-0.81 versus -0.76 for the most negatively correlated regions), with only the superior frontal gyrus, supramarginal gyrus, and superior temporal gyrus being still strongly correlated with t-UMAP0. The age dependency completely disappeared for the ADNI embedding both for t-UMAP0 and t-UMAP1. It is likely that, since ADNI’s age range is restricted, so is the cortical thickness loss due to aging. A possible other explanation comes from the fact that MCI and AD-related aging should not affect cortical thickness the same way as non-pathological aging. This way, the subject’s state shouldn’t be correlated with its age.

### Trajectory definition and analyses

From Figure 7c)and d) it is clear that MCI-MCI trajectories are different from MCI-AD. Both trajectory types have different attracting points and different averages, although it could be construed that most of these differences between trajectory types could be explained by the differences in the samples represented in each type. The ADNI embedding displays similar properties, as exemplified by differences in the average trajectories between MCI-MCI and MCI-AD in Figure 7d). For this embedding, since none of the axis are correlated with age, the differences between both groups can only be explained by an underlying property of the embedding. On the other hand, the average trajectories have a smaller norm overall. Scans coming from a same individual are usually grouped together in the same cluster.

### Clusters

Some interesting patterns emerge in the atlas shown in Figure 8a). First of all, there is a strong relationship between the final cluster and AD diagnosis. Indeed, trajectories with the most subjects show a trend where subjects with a final scan in cluster 0 have a low (*∼* 17%) probability of developing AD and the probability goes up with the index of the final cluster : cluster 1 *∼*30%, cluster 2 *∼*38% and cluster 3 *∼*62%. Moreover, the longer a subject stays in the cluster the stronger is this trend. For example, the 3-step trajectory “0-0-0” has a similar probability than its homologous 1-step trajectory “0”, while “1-1-1” shows a *∼*2% increase in probability when compared to “1”, “2-2-2” a *∼*10% increase when compared to “2” and “3-3-3” a *∼*15% increase when compared to “3”. Since age is distributed similarly between AD and MCI-MCI subjects, there seems to be a clear relationship between cluster 3 and the diagnosis of AD. Moreover, trajectories with an intermediary data point in cluster 3 and another in cluster 0, 1 or 2 afterwards, have a significantly lower probability to describe AD. This phenomenon is directly related to our observations that MCI-MCI subjects in cluster 3 go towards the other clusters on average, while AD subjects stay in cluster 3. From this figure we can then conclude the clusters and trajectories are correlated with certain diagnosis.

The ADNI embedding in Figure 8b) shows very similar trends. We already know that the first and last cluster are respectively associated with the lowest and highest AD probability. The behavior at the center is much different for the ADNI embedding, since the probability doesn’t seem to augment the closer you move to the right of the figure. Indeed we saw that cluster 1 (38%-41%) has a higher probability than cluster 2 (28%-31%). This discrepancy can be explained by the fact that clusters 1 and 2 are overlapping on the t-UMAP0 (see Figure 7) as compared to the NOMIS embedding where clusters mostly encompass a different range on the axis. Additionally, from Figure 5c) we observed that AD subjects seems to prefer lower values of the t-UMAP1 axis, which also supports the fact that cluster 1 is associated with a higher probability developing AD. From this, both embedding have prediction potential based on their clustering.

The predictive analysis shown in Figure 9 corroborates the previous observations. The AUC shows that trajectories, based on cortical thickness only, are able to classify subjects between MCI-MCIs and MCI-ADs in more than 72% of cases for the best models (6 clusters) on the test set. Even though the AUC is higher on average for trajectories of length 1 on the validation set, the models with the highest predictive power was, for both embedding with length of 3. For every model of the NOMIS embedding, the essential information necessary for classification seems to be located in the last cluster of the trajectory, since longer trajectory length reduces the accuracy and enhances the standard deviation on the validation set. For the ADNI embedding the predictive power on the validation set is more divided between length of one and two, with the model with four clusters having a higher AUC with length of two. On the test set, the ADNI embedding manages to outperform the NOMIS embedding with a respective AUC of 0.80 and 0.72. The embedding trained directly on the ADNI space set seems better suited for predicting decline than the NOMIS embedding, even though the ladder has a strong correlation with participants age. The hypothesis that an embedding representing normal aging would help differentiate participants on the AD spectrum is then incorrect.

## Conclusion

This study explored the visualization and statistical analysis of morphometric aging trajectories using UMAP, a novel dimensionality reduction technique, for subjects with AD or a MCI. We demonstrated the potential advantages of using this approach. We showed that using this method leads to an embedding space in which subjects exhibit trajectories between clusters that are significantly different depending on their remaining cognitively stable or declining to AD. Our results thus indicate that UMAP is a new and effective tool that not only can visualize aging trajectories, but also accurately differentiate between MCI-AD and MCI-MCI trajectories. To further improve the accuracy of the model, it would be of interest to perform a UMAP-based analysis using only the thickness of cortical regions related to AD^7,33,50^ and other data related to AD biomarkers such as amyloid beta concentration^51,52^, cognitive score^53^ and cognitive reserve^54^.

## Data Availability

All the datasets used in this manuscript are available online upon request to corresponding owner.

http://www.birncommunity.org/resources/data/

https://aibl.csiro.au/

http://www.mrc-cbu.cam.ac.uk/datasets/camcan/

http://www.cima-q.ca

https://www.alz.washington.edu/

http://www.oasis-brains.org/

http://brain-development.org/ixi-dataset/

https://www.nitrc.org/projects/multimodal

http://miriad.drc.ion.ucl.ac.uk

## Data availability

We are not the owners of the data used in this study, but all the databases are publicly available upon request. The links to the different databases and the instructions to obtain the datasets, can be found here :

Autism Brain Imaging Data Exchange (ABIDE): http://fcon_1000.projects.nitrc.org/indi/abide/

Alzheimer’s Disease Neuroimaging Initiative (ADNI): http://adni.loni.usc.edu/

Australian Imaging Biomarkers and Lifestyle flagship study of ageing (AIBL) : https://aibl.csiro.au/

Berlin Mind and Brain (Margulies, Villringer) CoRR sample (BMB): http://fcon_1000.projects.nitrc.org/indi/CoRR/html/bmb_1.html

Cambridge Centre for Ageing and Neuroscience (CamCAN): http://www.mrc-cbu.cam.ac.uk/datasets/camcan/

Center of Biomedical Research Excellence (COBRE): http://fcon_1000.projects.nitrc.org/indi/retro/cobre.html

Cleveland Clinic (Cleveland CCF): http://fcon_1000.projects.nitrc.org/indi/retro/ClevelandCCF.html

Consortium for the Early Identification of Alzheimer’s Disease (CIMA-Q): http://www.cima-q.ca

Dallas Lifespan Brain Study (DLBS): http://fcon_1000.projects.nitrc.org/indi/retro/dlbs.html

FIND lab sample: http://fcon_1000.projects.nitrc.org/indi/retro/find_stanford.html

Functional Biomedical Informatics Research Network (FBIRN): http://www.birncommunity.org/resources/data/

Lifespan Human Connectome Project in Aging (HCP-Aging): http://dx.doi.org/10.15154/1520138

International Consortium for Brain Mapping (ICBM): https://ida.loni.usc.edu/login.jsp?project=ICBM

Information eXtraction from Images (IXI): http://brain-development.org/ixi-dataset/

F.M. Kirby Research Center neuroimaging reproducibility data (KIRBY-21): https://www.nitrc.org/projects/multimodal

Minimal Interval Resonance Imaging in Alzheimer’s Disease (MIRIAD): http://miriad.drc.ion.ucl.ac.uk

National Alzheimer’s Coordinating Center (NACC): https://www.alz.washington.edu/

National Database for Autism Research (NDAR): http://nda.nih.gov

Nathan Kline Institute Rockland (NKI-R) sample (NKI-RS): http://fcon_1000.projects.nitrc.org/indi/pro/nki.html

NKI-R enhanced Sample (NKI-RES): http://fcon_1000.projects.nitrc.org/indi/enhanced/

Open access series of imaging studies (OASIS): http://www.oasis-brains.org/

POWER: http://fcon_1000.projects.nitrc.org/indi/retro/Power2012.html

Parkinson’s Progression Markers Initiative (PPMI): http://www.ppmi-info.org

Southwest University Adult Lifespan Dataset (SALD): http://fcon_1000.projects.nitrc.org/indi/retro/sald.html

University of Wisconsin, Madison (Birn, Prabhakaran, Meyerand) CoRR sample (UWM): http://fcon_1000.projects.nitrc.org/indi/CoRR/html/uwm_1.html

Wayne State EF Dataset: http://fcon_1000.projects.nitrc.org/indi/retro/wayne_EF.html

Yale Low-Resolution Controls Dataset: http://fcon_1000.projects.nitrc.org/indi/retro/yale_lowres.html

## 1 Acknowledgements

This work was supported by the Natural Sciences and Engineering Research Council of Canada (P.D., N.D), and the Sentinelle Nord program of Université Laval, funded by the Canada First Research Excellence Fund (P.D., N.D.). We acknowledge Calcul Québec and the Digital Research Alliance of Canada for their technical support and computing infrastructures.

This study comprises multiple samples of healthy individuals. We wish to thank all principal investigators who collected these datasets and agreed to let them accessible.

Autism Brain Imaging Data Exchange (ABIDE): Primary support for the work by Adriana Di Martino was provided by the NIMH (K23MH087770) and the Leon Levy Foundation. Primary support for the work by Michael P. Milham and the INDI team was provided by gifts from Joseph P. Healy and the Stavros Niarchos Foundation to the Child Mind Institute, as well as by an NIMH award to MPM (R03MH096321). http://fcon_1000.projects.nitrc.org/indi/abide/

Data collection and sharing for this project was funded by the Alzheimer’s Disease Neuroimaging Initiative (ADNI) (National Institutes of Health Grant U01 AG024904) and DOD ADNI (Department of Defense award number W81XWH-12-2-0012). ADNI is funded by the National Institute on Aging, the National Institute of Biomedical Imaging and Bioengineering, and through generous contributions from the following: AbbVie, Alzheimer’s Association; Alzheimer’s Drug Discovery Foundation; Araclon Biotech; BioClinica, Inc.; Biogen; Bristol-Myers Squibb Company; CereSpir, Inc.; Cogstate; Eisai Inc.; Elan Pharmaceuticals, Inc.; Eli Lilly and Company; EuroImmun; F. Hoffmann-La Roche Ltd and its affiliated company Genentech, Inc.; Fujirebio; GE Healthcare; IXICO Ltd.; Janssen Alzheimer Immunotherapy Research & Development, LLC.; Johnson & Johnson Pharmaceutical Research & Development LLC.; Lumosity; Lundbeck; Merck & Co., Inc.; Meso Scale Diagnostics, LLC.; NeuroRx Research; Neurotrack Technologies; Novartis Pharmaceuticals Corporation; Pfizer Inc.; Piramal Imaging; Servier; Takeda Pharmaceutical Company; and Transition Therapeutics. The Canadian Institutes of Health Research is providing funds to support ADNI clinical sites in Canada. Private sector contributions are facilitated by the Foundation for the National Institutes of Health (www.fnih.org). The grantee organization is the Northern California Institute for Research and Education, and the study is coordinated by the Alzheimer’s Therapeutic Research Institute at the University of Southern California. ADNI data are disseminated by the Laboratory for Neuro Imaging at the University of Southern California. http://adni.loni.usc.edu/. Data used in preparation of this article were obtained from the Alzheimer’s Disease Neuroimaging Initiative (ADNI) database (adni.loni.usc.edu). As such, the investigators within the ADNI contributed to the design and implementation of ADNI and/or provided data but did not participate in analysis or writing of this report. A complete listing of ADNI investigators can be found at [this link](https://adni.loni.usc.edu/wp-content/uploads/how_to_apply/ADNI_Acknowledgement_List.pdf)

Australian Imaging Biomarkers and Lifestyle flagship study of ageing (AIBL): Part of the data used in this study was obtained from the Australian Imaging Biomarkers and Lifestyle flagship study of ageing (AIBL) funded by the Commonwealth Scientific and Industrial Research Organisation (CSIRO) which was made available at the ADNI database (www.loni.usc.edu/ADNI). The AIBL researchers contributed data but did not participate in analysis or writing of this report. AIBL researchers are listed at www.aibl.csiro.au

Berlin Mind and Brain (Margulies, Villringer) CoRR sample (BMB). Zuo, X.N., et al. (2014). An open science resource for establishing reliability and reproducibility in functional connectomics. Scientific data, 1, 140049. doi: 10.1038/sdata.2014.49. http://fcon_1000.projects.nitrc.org/indi/CoRR/html/bmb_1.html

Cambridge Centre for Ageing and Neuroscience (CamCAN): CamCAN funding was provided by the UK Biotechnology and Biological Sciences Research Council (grant number BB/H008217/1), together with support from the UK Medical Research Council and University of Cambridge, UK. http://www.mrc-cbu.cam.ac.uk/datasets/camcan/

Center of Biomedical Research Excellence (COBRE): The imaging data and phenotypic information was collected and shared by the Mind Research Network and the University of New Mexico funded by a National Institute of Health COBRE: 1P20RR021938-01A2. http://fcon_1000.projects.nitrc.org/indi/retro/cobre.html

Cleveland Clinic (Cleveland CCF): Funded by the National Multiple Sclerosis Society. http://fcon_1000.projects.nitrc.org/indi/retro/ClevelandCCF.html

Consortium for the Early Identification of Alzheimer’s Disease (CIMA-Q): Part of the data used in this article were obtained from the Consortium pour l’identification précoce de la maladie Alzheimer - Québec (CIMA-Q). As such, the investigators within the CIMA-Q contributed to the design, the implementation, the acquisition of clinical, cognitive, and neuroimaging data and biological samples. A list of the CIMA-Q investigators is available on http://www.cima-q.ca. CIMA-Q was funded in 2013 with a $2,500,000 grant from the Fonds d’Innovation Pfizer - Fond de Recherche Québec – Santé sur la maladie d’Alzheimer et les maladies apparentées.

Dallas Lifespan Brain Study (DLBS): This study is supported by the Center for Vital Longevity, the University of Texas at Dallas, the University of Texas Southwestern Medical Center, the National Institutes of Health and Aging, AVID Radiopharmaceuticals, the Aging Mind Foundation and the Alzheimer’s Association. http://fcon_1000.projects.nitrc.org/indi/retro/dlbs.html

FIND lab sample. Funded by the Dana Foundation; John Douglas French Alzheimer’s Foundation; National Institutes of Health (AT005733, HD059205,HD057610, NS073498, NS058899). http://fcon_1000.projects.nitrc.org/indi/retro/find_stanford.html

Functional Biomedical Informatics Research Network (FBIRN): Provided by the Biomedical Informatics Research Network under the following support: U24-RR021992, by the National Center for Research Resources at the National Institutes of Health, U.S.A. http://www.birncommunity.org/resources/data/

Lifespan Human Connectome Project in Aging (HCP-Aging): HCP-Aging data were obtained from the National Institute of Mental Health (NIMH) Data Archive (NDA). NDA is a collaborative informatics system created by the National Institutes of Health to provide a national resource to support and accelerate research in mental health. Dataset identifier: http://dx.doi.org/10.15154/1520138. This manuscript reflects the views of the authors and may not reflect the opinions or views of the NIH or of the Submitters submitting original data to NDA. http://nda.nih.gov

International Consortium for Brain Mapping (ICBM). The ICBM (Principal Investigator: John Mazziotta, MD, PhD) was funded was provided by the National Institute of Biomedical Imaging and BioEngineering. ICBM is the result of efforts of co-investigators from UCLA, Montreal Neurologic Institute, University of Texas at San Antonio, and the Institute of Medicine, Juelich/Heinrich Heine University - Germany.” https://ida.loni.usc.edu/login.jsp?project=ICBM

Information eXtraction from Images (IXI): Data collected as part of the project EPSRC GR/S21533/02 - http://brain-development.org/ixi-dataset/

F.M. Kirby Research Center neuroimaging reproducibility data (KIRBY-21). Landman, B.A. et al. “Multi-Parametric Neuroimaging Reproducibility: A 3T Resource Study”, NeuroImage. (2010) NIHMS/PMC:252138 doi:10.1016/j.neuroimage.2010.11.047 https://www.nitrc.org/projects/multimodal

Minimal Interval Resonance Imaging in Alzheimer’s Disease (MIRIAD): The MIRIAD investigators did not participate in analysis or writing of this report. The MIRIAD dataset is made available through the support of the UK Alzheimer’s Society (RF116). The original data collection was funded through an unrestricted educational grant from GlaxoSmithKline (6GKC). http://miriad.drc.ion.ucl.ac.uk

National Alzheimer’s Coordinating Center (NACC): The NACC database is funded by NIA/NIH Grant U01 AG016976. NACC data are contributed by the NIA-funded ADCs: P30 AG019610 (PI Eric Reiman, MD), P30 AG013846 (PI Neil Kowall, MD), P30 AG062428-01 (PI James Leverenz, MD) P50 AG008702 (PI Scott Small, MD), P50 AG025688 (PI Allan Levey, MD, PhD), P50 AG047266 (PI Todd Golde, MD, PhD), P30 AG010133 (PI Andrew Saykin, PsyD), P50 AG005146 (PI Marilyn Albert, PhD), P30 AG062421-01 (PI Bradley Hyman, MD, PhD), P30 AG062422-01 (PI Ronald Petersen, MD, PhD), P50 AG005138 (PI Mary Sano, PhD), P30 AG008051 (PI Thomas Wisniewski, MD), P30 AG013854 (PI Robert Vassar, PhD), P30 AG008017 (PI Jeffrey Kaye, MD), P30 AG010161 (PI David Bennett, MD), P50 AG047366 (PI Victor Henderson, MD, MS), P30 AG010129 (PI Charles DeCarli, MD), P50 AG016573 (PI Frank LaFerla, PhD), P30 AG062429-01(PI James Brewer, MD, PhD), P50 AG023501 (PI Bruce Miller, MD), P30 AG035982 (PI Russell Swerdlow, MD), P30 AG028383 (PI Linda Van Eldik, PhD), P30 AG053760 (PI Henry Paulson, MD, PhD), P30 AG010124 (PI John Trojanowski, MD, PhD), P50 AG005133 (PI Oscar Lopez, MD), P50 AG005142 (PI Helena Chui, MD), P30 AG012300 (PI Roger Rosenberg, MD), P30 AG049638 (PI Suzanne Craft, PhD), P50 AG005136 (PI Thomas Grabowski, MD), P30 AG062715-01 (PI Sanjay Asthana, MD, FRCP), P50 AG005681 (PI John Morris, MD), P50 AG047270 (PI Stephen Strittmatter, MD, PhD). https://www.alz.washington.edu/

National Database for Autism Research (NDAR): Data were obtained from the National Institute of Mental Health (NIMH) Data Archive (NDA). NDA is a collaborative informatics system created by the National Institutes of Health to provide a national resource to support and accelerate research in mental health. Dataset identifier: http://dx.doi.org/10.15154/1520138. This manuscript reflects the views of the authors and may not reflect the opinions or views of the NIH or of the Submitters submitting original data to NDA. http://nda.nih.gov

Nathan Kline Institute Rockland (NKI-R) sample (NKI-RS) and Enhanced Sample (NKI-RES): Principal support for the NKI-RES project is provided by the NIMH BRAINS R01MH094639-01. Funding for key personnel also provided in part by the New York State Office of Mental Health and Research Foundation for Mental Hygiene. Funding for the decompression and augmentation of administrative and phenotypic protocols provided by a grant from the Child Mind Institute (1FDN2012-1). Additional personnel support provided by the Center for the Developing Brain at the Child Mind Institute, as well as NIMH R01MH081218, R01MH083246, and R21MH084126. Project support also provided by the NKI Center for Advanced Brain Imaging (CABI), the Brain Research Foundation, the Stavros Niarchos Foundation and the NIH P50 MH086385-S1 (NKI-RS). http://fcon_1000.projects.nitrc.org/indi/pro/nki.html http://fcon_1000.projects.nitrc.org/indi/enhanced/

Open access series of imaging studies (OASIS): The OASIS project was funded by grants P50 AG05681, P01 AG03991, R01 AG021910, P50 MH071616, U24 RR021382, and R01 MH56584. http://www.oasis-brains.org/

POWER: This database was supported by NIH R21NS061144 R01NS32979 R01HD057076 U54MH091657 K23DC006638 P50 MH71616 P60 DK020579-31, McDonnell Foundation Collaborative Action Award, NSF IGERT DGE-0548890, Simon’s Foundation Autism Research Initiative grant, Burroughs Wellcome Fund, Charles A. Dana Foundation, Brooks Family Fund, Tourette Syndrome Association, Barnes-Jewish Hospital Foundation, McDonnell Center for Systems Neuroscience, Alvin J. Siteman Cancer Center, American Hearing Research Foundation grant, Diabetes Research and Training Center at Washington University grant. http://fcon_1000.projects.nitrc.org/indi/retro/Power2012.html

Parkinson’s Progression Markers Initiative (PPMI): PPMI – a public-private partnership – is funded by the Michael J. Fox Foundation for Parkinson’s Research and funding partners, including Abbvie, Allergan, Amathus, Avid Radiopharmaceuticals, Biogen Idec, BioLegend, Bristol-Myers, Celgene, Cenali, Covance, GE Healthcare, Genentech, GlaxoSmithKline, Glolub Capital, Handl Therapeutics, Insitro, Janssen Neuroscience, Eli Lilly and Company, Lundbeck, Merck, Meso Scale Discovery, Neurocrine, Pfizer, Piramal, Prevail, Roche, Sanofi Genzyme, Servier, Takeda, Teva, UCB, Verily, and Voyager Therapeutics. See http://www.ppmi-info.org for further details.

Southwest University Adult Lifespan Dataset (SALD): SALD was supported by the National Natural Science Foundation of China (31470981; 31571137; 31500885), National Outstanding young people plan, the Program for the Top Young Talents by Chongqing, the Fundamental Research Funds for the Central Universities (SWU1509383,SWU1509451,SWU1609177), Natural Science Foundation of Chongqing (cstc2015jcyjA10106), Fok Ying Tung Education Foundation (151023), General Financial Grant from the China Postdoctoral Science Foundation (2015M572423, 2015M580767), Special Funds from the Chongqing Postdoctoral Science Foundation (Xm2015037, Xm2016044), Key research for Humanities and social sciences of Ministry of Education (14JJD880009). http://fcon_1000.projects.nitrc.org/indi/retro/sald.html

University of Wisconsin, Madison (Birn, Prabhakaran, Meyerand) CoRR sample (UWM): Zuo, X.N., et al. (2014). An open science resource for establishing reliability and reproducibility in functional connectomics. Scientific data, 1, 140049. doi: 10.1038/sdata.2014.49 http://fcon_1000.projects.nitrc.org/indi/CoRR/html/uwm_1.html

Wayne State EF Dataset: This dataset was supported by National Institute on Aging grants R01-AG011230, R37-AG011230, R03-AG024630 to Naftali Raz, Ph.D. http://fcon_1000.projects.nitrc.org/indi/retro/wayne_EF.html

Yale Low-Resolution Controls Dataset: Scheinost D, Tokoglu F, Shen X, Finn ES, Noble S, Papademetris X, Constable RT. Fluctuations in Global Brain Activity Are Associated With Changes in Whole-Brain Connectivity of Functional Networks. IEEE Trans Biomed Eng. 2016 Dec;63(12):2540-2549. Epub 2016 Aug 16. http://fcon_1000.projects.nitrc.org/indi/retro/yale_lowres.html

## Author contributions statement

N.D., P.D, and S.D. designed the project. O.P, R.L-C., and S.D. collected and prepared the data. R.L-C. wrote the codes and conducted the numerical experiments. All authors analyzed the results. R.L-C. wrote the main manuscript text while N.D., P.D, and S.D. revised, verified the results, and improved the writing of the manuscript.

