## Supplementary Table 1 for "Predicting cognitive decline in a low-dimensional representation of brain morphology"

| Cortical regions | Person correlation | Cortical regions | Person correlation |
|---|---|---|---|
| superiorfrontal_r | -0.876952 | parsorbitalis_r | -0.675987 |
| superiorfrontal_l | -0.875407 | lateralorbitofrontal_l | -0.66624 |
| supramarginal_l | -0.853448 | posteriorcingulate_l | -0.64067 |
| supramarginal_r | -0.834146 | lateralorbitofrontal_r | -0.633283 |
| inferiorparietal_l | -0.829533 | posteriorcingulate_r | -0.623178 |
| rostralmiddlefrontal_l | -0.826265 | lateraloccipital_l | -0.61807 |
| caudalmiddlefrontal_l | -0.824142 | lateraloccipital_r | -0.615145 |
| inferiorparietal_r | -0.820774 | fusiform_r | -0.605319 |
| superiortemporal_r | -0.806394 | inferiortemporal_r | -0.602292 |
| caudalmiddlefrontal_r | -0.805738 | caudalanteriorcingulate_l | -0.58978 |
| rostralmiddlefrontal_r | -0.80231 | inferiortemporal_l | -0.586643 |
| superiortemporal_l | -0.801841 | transversetemporal_r | -0.574015 |
| parstriangularis_l | -0.793562 | rostralanteriorcingulate_l | -0.573386 |
| middletemporal_r | -0.791966 | transversetemporal_l | -0.561767 |
| parsopercularis_r | -0.790692 | cuneus_l | -0.538545 |
| parstriangularis_r | -0.788787 | cuneus_r | -0.535733 |
| middletemporal_l | -0.785394 | medialorbitofrontal_r | -0.519666 |
| parsopercularis_l | -0.783172 | isthmuscingulate_l | -0.504857 |
| precuneus_l | -0.782538 | medialorbitofrontal_l | -0.501384 |
| precuneus_r | -0.771401 | isthmuscingulate_r | -0.489114 |
| precentral_l | -0.768512 | lingual_l | -0.434558 |
| precentral_r | -0.763119 | rostralanteriorcingulate_r | -0.423809 |
| superiorparietal_l | -0.740365 | lingual_r | -0.418618 |
| superiorparietal_r | -0.73864 | caudalanteriorcingulate_r | -0.407113 |
| postcentral_l | -0.716919 | pericalcarine_r | -0.343675 |
| insula_r | -0.711307 | parahippocampal_r | -0.330147 |
| paracentral_r | -0.699701 | pericalcarine_l | -0.318649 |
| postcentral_r | -0.69633 | parahippocampal_l | -0.304155 |
| paracentral_l | -0.689912 | entorhinal_l | -0.21535 |
| parsorbitalis_l | -0.686529 | entorhinal_r | -0.18102 |
| insula_l | -0.686017 | age | 0.649034 |
| fusiform_l | -0.682929 | | |

**Supplementary Table S1.** t-UMAP0's correlation with cortical regions and age. The Pearson correlation between the t-UMAP0 axis and the thickness of the different cortical regions used in the dimensional reduction and the subjects' age. These values were obtained with the UMAP parameters : `n_neighbors` = 20, `min_dist` = 0 and `random_state` = 42.
