## Supplementary Table 2 for "Predicting cognitive decline in a low-dimensional representation of brain morphology"

| Cortical regions | Person correlation | Cortical regions | Person correlation |
|---|---|---|---|
| pericalcarine_l | 0.552909 | superiortemporal_r | -0.0364093 |
| pericalcarine_r | 0.547725 | middletemporal_l | -0.0367981 |
| cuneus_r | 0.48084 | parstriangularis_l | -0.0373123 |
| lingual_r | 0.474045 | parsopercularis_l | -0.0509358 |
| lingual_l | 0.471035 | entorhinal_r | -0.0631519 |
| cuneus_l | 0.440937 | middletemporal_r | -0.0642912 |
| transversetemporal_l | 0.294846 | isthmuscingulate_r | -0.0678302 |
| transversetemporal_r | 0.274237 | rostralmiddlefrontal_l | -0.0854371 |
| lateraloccipital_r | 0.266643 | entorhinal_l | -0.0916308 |
| postcentral_r | 0.264153 | parsopercularis_r | -0.0959004 |
| postcentral_l | 0.259048 | parsorbitalis_l | -0.100174 |
| lateraloccipital_l | 0.251781 | parstriangularis_r | -0.10494 |
| superiorparietal_r | 0.225238 | superiorfrontal_l | -0.111455 |
| superiorparietal_l | 0.210182 | isthmuscingulate_l | -0.111712 |
| precentral_l | 0.193447 | inferiortemporal_r | -0.118378 |
| precentral_r | 0.180491 | parsorbitalis_r | -0.144352 |
| paracentral_r | 0.165205 | superiorfrontal_r | -0.147209 |
| precuneus_r | 0.144318 | rostralmiddlefrontal_r | -0.165768 |
| paracentral_l | 0.137474 | insula_r | -0.177589 |
| age | 0.128916 | insula_l | -0.180815 |
| precuneus_l | 0.118249 | posteriorcingulate_r | -0.185355 |
| parahippocampal_r | 0.078403 | posteriorcingulate_l | -0.190291 |
| fusiform_r | 0.0779755 | inferiortemporal_l | -0.195443 |
| inferiorparietal_r | 0.057548 | lateralorbitofrontal_l | -0.20209 |
| fusiform_l | 0.0571907 | medialorbitofrontal_l | -0.224996 |
| parahippocampal_l | 0.051324 | medialorbitofrontal_r | -0.228779 |
| inferiorparietal_l | 0.0422169 | caudalanteriorcingulate_l | -0.271913 |
| supramarginal_r | 0.0414675 | lateralorbitofrontal_r | -0.277769 |
| supramarginal_l | 0.030307 | caudalanteriorcingulate_r | -0.278458 |
| superiortemporal_l | 0.015318 | rostralanteriorcingulate_l | -0.289819 |
| caudalmiddlefrontal_l | -0.00400514 | rostralanteriorcingulate_r | -0.305279 |
| caudalmiddlefrontal_r | -0.0306759 | | |

**Supplementary Table S2.** t-UMAP1's correlation with cortical regions and age. The Pearson correlation between the t-UMAP1 axis and the thickness of the different cortical regions used in the dimensional reduction and the subjects' age. These values were obtained with the UMAP parameters : `n_neighbors` = 20, `min_dist` = 0 and `random_state` = 42.
