## Supplementary Table 3 for "Predicting cognitive decline in a low-dimensional representation of brain morphology"

| Cortical regions | Person correlation | Cortical regions | Person correlation |
|---|---|---|---|
| supramarginal_l | -0.805249 | paracentral_l | -0.60698 |
| superiortemporal_l | -0.760692 | superiorparietal_l | -0.602568 |
| superiorfrontal_r | -0.748668 | insula_l | -0.589436 |
| superiorfrontal_l | -0.745961 | isthmuscingulate_r | -0.586814 |
| supramarginal_r | -0.745148 | lateralorbitofrontal_l | -0.585709 |
| middletemporal_l | -0.73983 | lateralorbitofrontal_r | -0.58525 |
| inferiorparietal_r | -0.732523 | paracentral_r | -0.582005 |
| middletemporal_r | -0.732462 | transversetemporal_l | -0.578652 |
| inferiorparietal_l | -0.730992 | isthmuscingulate_l | -0.565313 |
| superiortemporal_r | -0.727618 | posteriorcingulate_l | -0.561167 |
| caudalmiddlefrontal_l | -0.715281 | transversetemporal_r | -0.555162 |
| rostralmiddlefrontal_l | -0.714318 | lingual_l | -0.552015 |
| precentral_l | -0.688223 | cuneus_r | -0.5468 |
| fusiform_l | -0.685649 | lingual_r | -0.543439 |
| inferiortemporal_l | -0.684909 | medialorbitofrontal_r | -0.538125 |
| postcentral_l | -0.679531 | parsorbitalis_r | -0.5379 |
| parstriangularis_l | -0.677658 | parahippocampal_r | -0.524067 |
| caudalmiddlefrontal_r | -0.674414 | parsorbitalis_l | -0.512752 |
| precentral_r | -0.673952 | cuneus_l | -0.499179 |
| precuneus_r | -0.669186 | posteriorcingulate_r | -0.494424 |
| rostralmiddlefrontal_r | -0.660783 | entorhinal_r | -0.489214 |
| precuneus_l | -0.654511 | entorhinal_l | -0.480666 |
| inferiortemporal_r | -0.651552 | rostralanteriorcingulate_l | -0.461693 |
| postcentral_r | -0.651014 | pericalcarine_r | -0.450832 |
| parsopercularis_r | -0.648596 | medialorbitofrontal_l | -0.448504 |
| parsopercularis_l | -0.64751 | pericalcarine_l | -0.446945 |
| lateraloccipital_l | -0.642215 | parahippocampal_l | -0.408416 |
| insula_r | -0.64139 | caudalanteriorcingulate_l | -0.398887 |
| fusiform_r | -0.636744 | rostralanteriorcingulate_r | -0.364156 |
| lateraloccipital_r | -0.62957 | caudalanteriorcingulate_r | -0.223265 |
| parstriangularis_r | -0.618905 | age | 0.407733 |
| superiorparietal_r | -0.609673 | | |

**Supplementary Table S3.** t-UMAP0's correlation with cortical regions and age for the ADNI embedding. The Pearson correlation between the t-UMAP0 axis and the thickness of the different cortical regions used in the dimensional reduction and the subjects' age. These values were obtained with the UMAP parameters : `n_neighbors` = 16, `min_dist` = 0 and `random_state` = 42.
