## Supplementary Table 4 for "Predicting cognitive decline in a low-dimensional representation of brain morphology"

| | | | |
|---|---|---|---|
| entorhinal_l | 0.537665 | parsopercularis_r | -0.0707972 |
| entorhinal_r | 0.518321 | posteriorcingulate_l | -0.0769428 |
| parahippocampal_r | 0.329262 | caudalanteriorcingulate_l | -0.084356 |
| parahippocampal_l | 0.328523 | parstriangularis_r | -0.104406 |
| inferiortemporal_r | 0.213144 | cuneus_r | -0.106049 |
| superiortemporal_r | 0.200402 | transversetemporal_l | -0.106363 |
| superiortemporal_l | 0.18822 | inferiorparietal_r | -0.108123 |
| inferiortemporal_l | 0.183617 | supramarginal_r | -0.113103 |
| middletemporal_r | 0.15229 | superiorfrontal_r | -0.118693 |
| fusiform_r | 0.149542 | rostralmiddlefrontal_r | -0.121148 |
| insula_l | 0.140429 | precuneus_r | -0.122006 |
| insula_r | 0.131936 | rostralmiddlefrontal_l | -0.125883 |
| fusiform_l | 0.127449 | precentral_r | -0.128905 |
| middletemporal_l | 0.123229 | cuneus_l | -0.131271 |
| medialorbitofrontal_r | 0.0737625 | supramarginal_l | -0.132471 |
| rostralanteriorcingulate_l | 0.0574475 | parsorbitalis_l | -0.138037 |
| lateralorbitofrontal_l | 0.0450584 | parsopercularis_l | -0.14382 |
| isthmuscingulate_l | 0.0210627 | posteriorcingulate_r | -0.153789 |
| medialorbitofrontal_l | 0.019158 | superiorparietal_r | -0.153912 |
| lateralorbitofrontal_r | 0.0166662 | precuneus_l | -0.155209 |
| pericalcarine_r | 0.00715549 | caudalanteriorcingulate_r | -0.161414 |
| age | 0.00322331 | paracentral_l | -0.164732 |
| pericalcarine_l | 0.00205908 | superiorparietal_l | -0.169337 |
| rostralanteriorcingulate_r | -0.00119598 | parstriangularis_l | -0.173961 |
| lingual_l | -0.018495 | superiorfrontal_l | -0.173991 |
| isthmuscingulate_r | -0.0297047 | caudalmiddlefrontal_l | -0.174653 |
| lateraloccipital_l | -0.0488989 | postcentral_r | -0.176028 |
| lateraloccipital_r | -0.0558424 | caudalmiddlefrontal_r | -0.180872 |
| transversetemporal_r | -0.0590221 | precentral_l | -0.189614 |
| parsorbitalis_r | -0.0665719 | postcentral_l | -0.21358 |
| inferiorparietal_l | -0.0684489 | paracentral_r | -0.218963 |
| lingual_r | -0.0685776 | | |

**Supplementary Table S4.** t-UMAP1's correlation with cortical regions and age for the ADNI embedding. The Pearson correlation between the t-UMAP0 axis and the thickness of the different cortical regions used in the dimensional reduction and the subjects' age. These values were obtained with the UMAP parameters : `n_neighbors` = 16, `min_dist` = 0 and `random_state` = 42.
