## Supplementary Table 5 for "Predicting cognitive decline in a low-dimensional representation of brain morphology"

| Diagnosis | At Baseline | At Last follow-up |
|---|---|---|
| CH | 526 | 477 |
| MCI | 828 | 537 |
| AD | 0 | 340 |

**Supplementary Table S5.** The distribution of diagnosis in the ADNI database at baseline and on the last available follow-up. These values were calculated from the raw data obtained.
