## Supplementary Table 6 for "Predicting cognitive decline in a low-dimensional representation of brain morphology"

| | | | | | | | |
|---|---|---|---|---|---|---|---|
| 002_S_0295 | 002_S_0413 | 002_S_0559 | 002_S_0619 | 002_S_0685 | 002_S_0729 | 002_S_0782 | 002_S_0816 |
| 002_S_0938 | 002_S_0954 | 002_S_0955 | 002_S_1018 | 002_S_1070 | 002_S_1155 | 002_S_1261 | 002_S_1268 |
| 002_S_1280 | 003_S_0907 | 003_S_0908 | 003_S_0931 | 003_S_0981 | 003_S_1021 | 003_S_1057 | 003_S_1059 |
| 003_S_1074 | 003_S_1122 | 003_S_1257 | 005_S_0221 | 005_S_0222 | 005_S_0223 | 005_S_0324 | 005_S_0448 |
| 005_S_0546 | 005_S_0553 | 005_S_0572 | 005_S_0602 | 005_S_0610 | 005_S_0814 | 005_S_0929 | 005_S_1341 |
| 006_S_0484 | 006_S_0498 | 006_S_0547 | 006_S_0653 | 006_S_0675 | 006_S_0681 | 006_S_0731 | 006_S_1130 |
| 007_S_0041 | 007_S_0068 | 007_S_0070 | 007_S_0101 | 007_S_0128 | 007_S_0249 | 007_S_0293 | 007_S_0316 |
| 007_S_0344 | 007_S_0414 | 007_S_0698 | 007_S_1206 | 007_S_1222 | 007_S_1248 | 007_S_1304 | 007_S_1339 |
| 009_S_0751 | 009_S_0842 | 009_S_0862 | 009_S_1030 | 009_S_1199 | 009_S_1334 | 009_S_1354 | 010_S_0067 |
| 010_S_0161 | 010_S_0419 | 010_S_0420 | 010_S_0422 | 010_S_0472 | 010_S_0786 | 010_S_0829 | 010_S_0904 |
| 011_S_0002 | 011_S_0003 | 011_S_0005 | 011_S_0008 | 011_S_0010 | 011_S_0016 | 011_S_0021 | 011_S_0022 |
| 011_S_0023 | 011_S_0053 | 011_S_0168 | 011_S_0183 | 011_S_0241 | 011_S_0326 | 011_S_0362 | 011_S_0856 |
| 011_S_0861 | 011_S_1080 | 011_S_1282 | 012_S_0634 | 012_S_0637 | 012_S_0689 | 012_S_0712 | 012_S_0720 |
| 012_S_0803 | 012_S_0917 | 012_S_0932 | 012_S_1009 | 012_S_1033 | 012_S_1133 | 012_S_1165 | 012_S_1175 |
| 012_S_1212 | 012_S_1292 | 012_S_1321 | 013_S_0240 | 013_S_0325 | 013_S_0502 | 013_S_0575 | 013_S_0592 |
| 013_S_0699 | 013_S_0860 | 013_S_0996 | 013_S_1035 | 013_S_1120 | 013_S_1161 | 013_S_1186 | 013_S_1205 |
| 013_S_1275 | 013_S_1276 | 014_S_0169 | 014_S_0328 | 014_S_0356 | 014_S_0519 | 014_S_0520 | 014_S_0548 |
| 014_S_0557 | 014_S_0558 | 014_S_0563 | 014_S_0658 | 014_S_1095 | 016_S_0354 | 016_S_0359 | 016_S_0538 |
| 016_S_0590 | 016_S_0702 | 016_S_0769 | 016_S_0991 | 016_S_1028 | 016_S_1092 | 016_S_1117 | 016_S_1121 |
| 016_S_1138 | 016_S_1149 | 016_S_1263 | 016_S_1326 | 018_S_0043 | 018_S_0057 | 018_S_0080 | 018_S_0087 |
| 018_S_0103 | 018_S_0142 | 018_S_0155 | 018_S_0286 | 018_S_0335 | 018_S_0369 | 018_S_0406 | 018_S_0425 |
| 018_S_0450 | 018_S_0633 | 018_S_0682 | 020_S_0097 | 020_S_0213 | 020_S_0883 | 020_S_0899 | 020_S_1288 |
| 021_S_0141 | 021_S_0159 | 021_S_0178 | 021_S_0231 | 021_S_0273 | 021_S_0276 | 021_S_0332 | 021_S_0337 |
| 021_S_0343 | 021_S_0424 | 021_S_0626 | 021_S_0642 | 021_S_0647 | 021_S_0753 | 021_S_0984 | 021_S_1109 |
| 022_S_0004 | 022_S_0007 | 022_S_0014 | 022_S_0044 | 022_S_0066 | 022_S_0096 | 022_S_0129 | 022_S_0130 |
| 022_S_0219 | 022_S_0543 | 022_S_0544 | 022_S_0750 | 022_S_0924 | 022_S_0961 | 022_S_1097 | 022_S_1351 |
| 022_S_1366 | 022_S_1394 | 023_S_0031 | 023_S_0042 | 023_S_0058 | 023_S_0061 | 023_S_0078 | 023_S_0081 |
| 023_S_0083 | 023_S_0084 | 023_S_0093 | 023_S_0126 | 023_S_0139 | 023_S_0217 | 023_S_0331 | 023_S_0376 |
| 023_S_0388 | 023_S_0604 | 023_S_0613 | 023_S_0625 | 023_S_0855 | 023_S_0887 | 023_S_0916 | 023_S_0926 |
| 023_S_0963 | 023_S_1046 | 023_S_1104 | 023_S_1126 | 023_S_1190 | 023_S_1247 | 023_S_1262 | 023_S_1289 |
| 023_S_1306 | 024_S_0985 | 024_S_1063 | 024_S_1171 | 024_S_1307 | 024_S_1393 | 024_S_1400 | 027_S_0074 |
| 027_S_0116 | 027_S_0118 | 027_S_0120 | 027_S_0179 | 027_S_0256 | 027_S_0307 | 027_S_0403 | 027_S_0404 |
| 027_S_0408 | 027_S_0417 | 027_S_0461 | 027_S_0485 | 027_S_0644 | 027_S_0835 | 027_S_0850 | 027_S_1045 |
| 027_S_1081 | 027_S_1082 | 027_S_1213 | 027_S_1254 | 027_S_1277 | 027_S_1385 | 027_S_1387 | 029_S_0824 |
| 029_S_0836 | 029_S_0843 | 029_S_0845 | 029_S_0866 | 029_S_0871 | 029_S_0878 | 029_S_0914 | 029_S_0999 |
| 029_S_1038 | 029_S_1056 | 029_S_1073 | 029_S_1184 | 029_S_1218 | 029_S_1318 | 029_S_1384 | 031_S_0294 |
| 031_S_0321 | 031_S_0351 | 031_S_0554 | 031_S_0568 | 031_S_0618 | 031_S_0821 | 031_S_0830 | 031_S_0867 |
| 031_S_1066 | 031_S_1209 | 032_S_0095 | 032_S_0147 | 032_S_0187 | 032_S_0214 | 032_S_0400 | 032_S_0479 |
| 032_S_0677 | 032_S_0718 | 032_S_0978 | 032_S_1037 | 032_S_1101 | 032_S_1169 | 033_S_0511 | 033_S_0513 |
| 033_S_0514 | 033_S_0516 | 033_S_0567 | 033_S_0723 | 033_S_0724 | 033_S_0725 | 033_S_0733 | 033_S_0734 |
| 033_S_0739 | 033_S_0741 | 033_S_0889 | 033_S_0906 | 033_S_0920 | 033_S_0922 | 033_S_0923 | 033_S_1016 |
| 033_S_1086 | 033_S_1098 | 033_S_1116 | 033_S_1279 | 033_S_1281 | 033_S_1283 | 033_S_1284 | 033_S_1285 |
| 033_S_1308 | 033_S_1309 | 035_S_0033 | 035_S_0048 | 035_S_0156 | 035_S_0204 | 035_S_0292 | 035_S_0341 |
| 035_S_0555 | 035_S_0997 | 036_S_0576 | 036_S_0577 | 036_S_0656 | 036_S_0672 | 036_S_0673 | 036_S_0748 |
| 036_S_0759 | 036_S_0760 | 036_S_0813 | 036_S_0869 | 036_S_0945 | 036_S_0976 | 036_S_1001 | 036_S_1023 |
| 036_S_1135 | 036_S_1240 | 037_S_0150 | 037_S_0182 | 037_S_0303 | 037_S_0327 | 037_S_0377 | 037_S_0454 |
| 037_S_0467 | 037_S_0501 | 037_S_0539 | 037_S_0552 | 037_S_0566 | 037_S_0588 | 037_S_0627 | 037_S_1078 |
| 037_S_1225 | 037_S_1421 | 041_S_0125 | 041_S_0262 | 041_S_0282 | 041_S_0314 | 041_S_0407 | 041_S_0446 |
| 041_S_0549 | 041_S_0598 | 041_S_0679 | 041_S_0721 | 041_S_0898 | 041_S_1002 | 041_S_1010 | 041_S_1260 |
| 041_S_1368 | 041_S_1391 | 041_S_1411 | 041_S_1412 | 041_S_1418 | 041_S_1420 | 041_S_1423 | 041_S_1425 |
| 041_S_1435 | 051_S_1040 | 051_S_1072 | 051_S_1123 | 051_S_1131 | 051_S_1331 | 051_S_1338 | 052_S_0671 |
| 052_S_0951 | 052_S_0952 | 052_S_0989 | 052_S_1054 | 052_S_1168 | 052_S_1250 | 052_S_1251 | 052_S_1346 |
| 052_S_1352 | 053_S_0389 | 053_S_0507 | 053_S_0621 | 053_S_0919 | 053_S_1044 | 057_S_0464 | 057_S_0474 |

**Supplementary Table S6.** List of ADNI subjects' ID used in this study.
