## Supplementary Table 7 for "Predicting cognitive decline in a low-dimensional representation of brain morphology"

| | | | | | | | |
|---|---|---|---|---|---|---|---|
| 057_S_0643 | 057_S_0779 | 057_S_0818 | 057_S_0839 | 057_S_0934 | 057_S_0941 | 057_S_0957 | 057_S_1007 |
| 057_S_1217 | 057_S_1265 | 057_S_1269 | 057_S_1371 | 057_S_1373 | 057_S_1379 | 062_S_0535 | 062_S_0578 |
| 062_S_0690 | 062_S_0730 | 062_S_0768 | 062_S_0793 | 062_S_1099 | 062_S_1182 | 062_S_1294 | 062_S_1299 |
| 067_S_0019 | 067_S_0029 | 067_S_0038 | 067_S_0045 | 067_S_0056 | 067_S_0059 | 067_S_0076 | 067_S_0077 |
| 067_S_0098 | 067_S_0110 | 067_S_0176 | 067_S_0177 | 067_S_0243 | 067_S_0257 | 067_S_0284 | 067_S_0290 |
| 067_S_0336 | 067_S_0607 | 067_S_0812 | 067_S_0828 | 067_S_1185 | 067_S_1253 | 068_S_0109 | 068_S_0127 |
| 068_S_0210 | 068_S_0401 | 068_S_0442 | 068_S_0473 | 068_S_0476 | 068_S_0478 | 068_S_0802 | 068_S_0872 |
| 068_S_1075 | 068_S_1191 | 072_S_0315 | 072_S_1211 | 072_S_1380 | 073_S_0089 | 073_S_0311 | 073_S_0312 |
| 073_S_0386 | 073_S_0445 | 073_S_0518 | 073_S_0565 | 073_S_0746 | 073_S_0909 | 073_S_1357 | 082_S_0304 |
| 082_S_0363 | 082_S_0469 | 082_S_0640 | 082_S_0641 | 082_S_0761 | 082_S_0832 | 082_S_0928 | 082_S_1079 |
| 082_S_1119 | 082_S_1256 | 082_S_1377 | 094_S_0434 | 094_S_0489 | 094_S_0526 | 094_S_0531 | 094_S_0692 |
| 094_S_0711 | 094_S_0921 | 094_S_1015 | 094_S_1027 | 094_S_1090 | 094_S_1102 | 094_S_1164 | 094_S_1188 |
| 094_S_1241 | 094_S_1267 | 094_S_1293 | 094_S_1314 | 094_S_1330 | 094_S_1397 | 094_S_1398 | 094_S_1402 |
| 094_S_1417 | 098_S_0149 | 098_S_0160 | 098_S_0171 | 098_S_0172 | 098_S_0269 | 098_S_0288 | 098_S_0667 |
| 098_S_0884 | 098_S_0896 | 099_S_0040 | 099_S_0051 | 099_S_0054 | 099_S_0060 | 099_S_0090 | 099_S_0111 |
| 099_S_0291 | 099_S_0352 | 099_S_0372 | 099_S_0470 | 099_S_0492 | 099_S_0533 | 099_S_0534 | 099_S_0551 |
| 099_S_0880 | 099_S_0958 | 099_S_1034 | 099_S_1144 | 100_S_0006 | 100_S_0015 | 100_S_0035 | 100_S_0047 |
| 100_S_0069 | 100_S_0190 | 100_S_0296 | 100_S_0743 | 100_S_0747 | 100_S_0892 | 100_S_0930 | 100_S_0995 |
| 100_S_1062 | 100_S_1113 | 100_S_1154 | 100_S_1226 | 100_S_1286 | 109_S_0777 | 109_S_0876 | 109_S_0950 |
| 109_S_0967 | 109_S_1013 | 109_S_1014 | 109_S_1114 | 109_S_1157 | 109_S_1183 | 109_S_1192 | 109_S_1343 |
| 114_S_0166 | 114_S_0173 | 114_S_0228 | 114_S_0374 | 114_S_0378 | 114_S_0410 | 114_S_0416 | 114_S_0458 |
| 114_S_0601 | 114_S_0979 | 114_S_1103 | 114_S_1106 | 114_S_1118 | 116_S_0360 | 116_S_0361 | 116_S_0370 |
| 116_S_0382 | 116_S_0392 | 116_S_0487 | 116_S_0648 | 116_S_0649 | 116_S_0657 | 116_S_0752 | 116_S_0834 |
| 116_S_0890 | 116_S_1083 | 116_S_1232 | 116_S_1243 | 116_S_1249 | 116_S_1271 | 116_S_1315 | 121_S_1322 |
| 121_S_1350 | 123_S_0050 | 123_S_0072 | 123_S_0088 | 123_S_0091 | 123_S_0094 | 123_S_0106 | 123_S_0108 |
| 123_S_0113 | 123_S_0162 | 123_S_0298 | 123_S_0390 | 123_S_1300 | 126_S_0405 | 126_S_0506 | 126_S_0605 |
| 126_S_0606 | 126_S_0680 | 126_S_0708 | 126_S_0709 | 126_S_0784 | 126_S_0865 | 126_S_0891 | 126_S_1077 |
| 126_S_1187 | 126_S_1221 | 126_S_1340 | 127_S_0112 | 127_S_0259 | 127_S_0260 | 127_S_0393 | 127_S_0394 |
| 127_S_0397 | 127_S_0431 | 127_S_0622 | 127_S_0684 | 127_S_0754 | 127_S_0844 | 127_S_0925 | 127_S_1032 |
| 127_S_1140 | 127_S_1210 | 127_S_1382 | 127_S_1419 | 127_S_1427 | 128_S_0135 | 128_S_0138 | 128_S_0167 |
| 128_S_0188 | 128_S_0200 | 128_S_0205 | 128_S_0216 | 128_S_0225 | 128_S_0227 | 128_S_0229 | 128_S_0230 |
| 128_S_0245 | 128_S_0258 | 128_S_0266 | 128_S_0272 | 128_S_0310 | 128_S_0500 | 128_S_0517 | 128_S_0522 |
| 128_S_0528 | 128_S_0545 | 128_S_0608 | 128_S_0611 | 128_S_0715 | 128_S_0740 | 128_S_0770 | 128_S_0863 |
| 128_S_0947 | 128_S_1043 | 128_S_1088 | 128_S_1148 | 128_S_1242 | 128_S_1406 | 128_S_1407 | 128_S_1408 |
| 128_S_1409 | 128_S_1430 | 129_S_0778 | 129_S_1204 | 129_S_1246 | 130_S_0102 | 130_S_0232 | 130_S_0285 |
| 130_S_0289 | 130_S_0423 | 130_S_0449 | 130_S_0505 | 130_S_0783 | 130_S_0886 | 130_S_0956 | 130_S_0969 |
| 130_S_1200 | 130_S_1201 | 130_S_1290 | 130_S_1337 | 131_S_0123 | 131_S_0319 | 131_S_0384 | 131_S_0409 |
| 131_S_0436 | 131_S_0441 | 131_S_0457 | 131_S_0497 | 131_S_0691 | 131_S_1301 | 131_S_1389 | 132_S_0339 |
| 132_S_0987 | 133_S_0433 | 133_S_0488 | 133_S_0493 | 133_S_0525 | 133_S_0629 | 133_S_0638 | 133_S_0727 |
| 133_S_0771 | 133_S_0792 | 133_S_0912 | 133_S_0913 | 133_S_1031 | 133_S_1055 | 133_S_1170 | 136_S_0086 |
| 136_S_0107 | 136_S_0184 | 136_S_0186 | 136_S_0194 | 136_S_0195 | 136_S_0196 | 136_S_0299 | 136_S_0300 |
| 136_S_0426 | 136_S_0429 | 136_S_0579 | 136_S_0695 | 136_S_0873 | 136_S_0874 | 136_S_1227 | 137_S_0158 |
| 137_S_0283 | 137_S_0301 | 137_S_0366 | 137_S_0438 | 137_S_0443 | 137_S_0459 | 137_S_0481 | 137_S_0631 |
| 137_S_0668 | 137_S_0669 | 137_S_0686 | 137_S_0722 | 137_S_0796 | 137_S_0800 | 137_S_0825 | 137_S_0841 |
| 137_S_0972 | 137_S_0973 | 137_S_0994 | 137_S_1041 | 137_S_1414 | 141_S_0696 | 141_S_0697 | 141_S_0717 |
| 141_S_0726 | 141_S_0767 | 141_S_0790 | 141_S_0810 | 141_S_0851 | 141_S_0852 | 141_S_0853 | 141_S_0915 |
| 141_S_0982 | 141_S_1004 | 141_S_1024 | 141_S_1051 | 141_S_1052 | 141_S_1094 | 141_S_1137 | 141_S_1152 |
| 141_S_1231 | 141_S_1244 | 141_S_1245 | 141_S_1255 | 141_S_1378 | 941_S_1194 | 941_S_1195 | 941_S_1197 |
| 941_S_1202 | 941_S_1203 | 941_S_1295 | 941_S_1311 | 941_S_1363 | 002_S_2010 | 002_S_2073 | 005_S_2390 |
| 007_S_2058 | 007_S_2106 | 007_S_2394 | 009_S_2208 | 009_S_2381 | 011_S_2274 | 013_S_2324 | 013_S_2389 |
| 014_S_2185 | 014_S_2308 | 016_S_2007 | 016_S_2031 | 018_S_0055 | 018_S_2133 | 018_S_2138 | 018_S_2155 |
| 018_S_2180 | 021_S_2077 | 021_S_2100 | 021_S_2124 | 021_S_2125 | 021_S_2142 | 021_S_2150 | 022_S_2087 |
| 022_S_2167 | 022_S_2263 | 022_S_2379 | 023_S_2068 | 024_S_2239 | 027_S_2183 | 027_S_2219 | 027_S_2245 |

**Supplementary Table S7.** List of ADNI subjects' ID used in this study (continuation).
