## Supplementary Table 8 for "Predicting cognitive decline in a low-dimensional representation of brain morphology"

| | | | | | | | |
|---|---|---|---|---|---|---|---|
| 027_S_2336 | 029_S_2376 | 029_S_2395 | 031_S_2018 | 031_S_2022 | 031_S_2233 | 032_S_2119 | 032_S_2240 |
| 032_S_2247 | 035_S_2061 | 035_S_2074 | 036_S_2378 | 036_S_2380 | 052_S_2249 | 053_S_2357 | 053_S_2396 |
| 057_S_2398 | 067_S_2195 | 067_S_2196 | 067_S_2301 | 067_S_2304 | 068_S_2168 | 068_S_2171 | 068_S_2184 |
| 068_S_2187 | 068_S_2193 | 068_S_2194 | 068_S_2248 | 068_S_2315 | 068_S_2316 | 072_S_2026 | 072_S_2027 |
| 072_S_2037 | 072_S_2072 | 072_S_2083 | 072_S_2093 | 072_S_2116 | 072_S_2164 | 073_S_2153 | 073_S_2182 |
| 073_S_2190 | 073_S_2191 | 073_S_2225 | 073_S_2264 | 082_S_2099 | 082_S_2121 | 082_S_2307 | 094_S_2201 |
| 094_S_2216 | 094_S_2238 | 094_S_2367 | 098_S_2047 | 098_S_2052 | 098_S_2079 | 099_S_2042 | 099_S_2063 |
| 099_S_2146 | 099_S_2205 | 114_S_2392 | 123_S_2055 | 123_S_2363 | 126_S_2360 | 126_S_2405 | 126_S_2407 |
| 127_S_2213 | 127_S_2234 | 128_S_2002 | 128_S_2036 | 128_S_2045 | 128_S_2123 | 129_S_2332 | 129_S_2347 |
| 130_S_2373 | 130_S_2403 | 141_S_2210 | 141_S_2333 | 153_S_2109 | 153_S_2148 | 002_S_2043 | 003_S_2374 |
| 016_S_2284 | 109_S_2200 | 128_S_2130 | 128_S_2151 | 128_S_2220 | 941_S_2060 | 035_S_2199 | 072_S_2070 |
| 109_S_2237 | 109_S_2278 | 128_S_2003 | 128_S_2011 | 128_S_2057 | 128_S_2314 | 002_S_4171 | 002_S_4213 |
| 002_S_4219 | 002_S_4225 | 002_S_4229 | 002_S_4237 | 002_S_4251 | 002_S_4262 | 002_S_4264 | 002_S_4270 |
| 002_S_4447 | 002_S_4473 | 002_S_4521 | 002_S_4654 | 002_S_4746 | 002_S_4799 | 002_S_5018 | 002_S_5178 |
| 002_S_5230 | 002_S_5256 | 003_S_4081 | 003_S_4119 | 003_S_4152 | 003_S_4288 | 003_S_4350 | 003_S_4354 |
| 003_S_4373 | 003_S_4441 | 003_S_4524 | 003_S_4555 | 003_S_4644 | 003_S_4872 | 003_S_4892 | 003_S_4900 |
| 003_S_5130 | 003_S_5150 | 003_S_5154 | 003_S_5165 | 003_S_5187 | 003_S_5209 | 005_S_4168 | 005_S_4185 |
| 005_S_4707 | 005_S_4910 | 005_S_5038 | 005_S_5119 | 006_S_4150 | 006_S_4153 | 006_S_4192 | 006_S_4346 |
| 006_S_4357 | 006_S_4363 | 006_S_4449 | 006_S_4485 | 006_S_4515 | 006_S_4546 | 006_S_4679 | 006_S_4713 |
| 006_S_4867 | 006_S_4960 | 006_S_5153 | 007_S_4272 | 007_S_4387 | 007_S_4467 | 007_S_4488 | 007_S_4516 |
| 007_S_4568 | 007_S_4611 | 007_S_4620 | 007_S_4637 | 007_S_5196 | 007_S_5265 | 009_S_4324 | 009_S_4337 |
| 009_S_4359 | 009_S_4388 | 009_S_4530 | 009_S_4543 | 009_S_4564 | 009_S_4612 | 009_S_4741 | 009_S_4814 |
| 009_S_4903 | 009_S_4958 | 009_S_5000 | 009_S_5027 | 009_S_5037 | 009_S_5125 | 009_S_5147 | 009_S_5176 |
| 009_S_5224 | 009_S_5252 | 010_S_4345 | 010_S_4442 | 011_S_4075 | 011_S_4105 | 011_S_4120 | 011_S_4222 |
| 011_S_4235 | 011_S_4278 | 011_S_4366 | 011_S_4547 | 011_S_4827 | 011_S_4845 | 011_S_4893 | 011_S_4906 |
| 011_S_4912 | 011_S_4949 | 012_S_4012 | 012_S_4026 | 012_S_4094 | 012_S_4188 | 012_S_4643 | 012_S_5121 |
| 012_S_5157 | 013_S_4268 | 013_S_4395 | 013_S_4579 | 013_S_4580 | 013_S_4595 | 013_S_4616 | 013_S_4791 |
| 013_S_4917 | 013_S_4985 | 013_S_5071 | 013_S_5137 | 013_S_5171 | 014_S_4039 | 014_S_4058 | 014_S_4079 |
| 014_S_4080 | 014_S_4093 | 014_S_4263 | 014_S_4328 | 014_S_4401 | 014_S_4576 | 014_S_4577 | 014_S_4615 |
| 014_S_4668 | 016_S_4009 | 016_S_4097 | 016_S_4121 | 016_S_4353 | 016_S_4575 | 016_S_4583 | 016_S_4584 |
| 016_S_4591 | 016_S_4601 | 016_S_4638 | 016_S_4646 | 016_S_4688 | 016_S_4887 | 016_S_4902 | 016_S_4951 |
| 016_S_4952 | 016_S_4963 | 016_S_5007 | 016_S_5031 | 016_S_5032 | 016_S_5057 | 018_S_4257 | 018_S_4313 |
| 018_S_4349 | 018_S_4399 | 018_S_4400 | 018_S_4597 | 018_S_4696 | 018_S_4733 | 018_S_4809 | 018_S_4868 |
| 018_S_4889 | 018_S_5240 | 018_S_5250 | 018_S_5262 | 019_S_4252 | 019_S_4285 | 019_S_4293 | 019_S_4367 |
| 019_S_4477 | 019_S_4548 | 019_S_4549 | 019_S_4680 | 019_S_4835 | 019_S_5012 | 019_S_5019 | 019_S_5242 |
| 020_S_4920 | 020_S_5140 | 020_S_5203 | 021_S_4245 | 021_S_4254 | 021_S_4276 | 021_S_4335 | 021_S_4402 |
| 021_S_4419 | 021_S_4421 | 021_S_4558 | 021_S_4633 | 021_S_4659 | 021_S_4718 | 021_S_4744 | 021_S_4857 |
| 021_S_4924 | 021_S_5099 | 021_S_5129 | 021_S_5177 | 021_S_5194 | 021_S_5236 | 021_S_5237 | 022_S_4173 |
| 022_S_4266 | 022_S_4291 | 022_S_4320 | 022_S_4444 | 022_S_4805 | 022_S_4922 | 022_S_5004 | 023_S_4020 |
| 023_S_4034 | 023_S_4035 | 023_S_4115 | 023_S_4122 | 023_S_4164 | 023_S_4241 | 023_S_4243 | 023_S_4448 |
| 023_S_4501 | 023_S_4502 | 023_S_4796 | 023_S_5241 | 024_S_4084 | 024_S_4158 | 024_S_4169 | 024_S_4186 |
| 024_S_4223 | 024_S_4280 | 024_S_4392 | 024_S_4622 | 024_S_4674 | 024_S_4905 | 024_S_5054 | 024_S_5290 |
| 027_S_4729 | 027_S_4757 | 027_S_4801 | 027_S_4802 | 027_S_4804 | 027_S_4869 | 027_S_4873 | 027_S_4919 |
| 027_S_4926 | 027_S_4936 | 027_S_4938 | 027_S_4943 | 027_S_4955 | 027_S_4962 | 027_S_4964 | 027_S_4966 |
| 027_S_5079 | 027_S_5083 | 027_S_5093 | 027_S_5109 | 027_S_5110 | 027_S_5118 | 027_S_5127 | 027_S_5169 |
| 027_S_5170 | 027_S_5277 | 027_S_5288 | 029_S_4279 | 029_S_4307 | 029_S_4327 | 029_S_4384 | 029_S_4385 |
| 029_S_4585 | 029_S_4652 | 029_S_5135 | 029_S_5158 | 029_S_5166 | 029_S_5219 | 031_S_4005 | 031_S_4021 |
| 031_S_4024 | 031_S_4029 | 031_S_4032 | 031_S_4042 | 031_S_4149 | 031_S_4194 | 031_S_4203 | 031_S_4218 |
| 031_S_4474 | 031_S_4476 | 031_S_4496 | 031_S_4590 | 031_S_4947 | 032_S_4277 | 032_S_4348 | 032_S_4386 |
| 032_S_4429 | 032_S_4755 | 032_S_4823 | 032_S_4921 | 032_S_5263 | 032_S_5289 | 033_S_4176 | 033_S_4177 |
| 033_S_4179 | 033_S_4505 | 033_S_4508 | 033_S_5013 | 033_S_5017 | 033_S_5087 | 033_S_5198 | 033_S_5235 |
| 033_S_5259 | 035_S_4082 | 035_S_4085 | 035_S_4114 | 035_S_4256 | 035_S_4414 | 035_S_4464 | 035_S_4582 |
| 035_S_4783 | 035_S_4784 | 035_S_4785 | 036_S_4389 | 036_S_4430 | 036_S_4491 | 036_S_4538 | 036_S_4562 |

**Supplementary Table S8.** List of ADNI subjects' ID used in this study (continuation).
