## Supplementary Table 9 for "Predicting cognitive decline in a low-dimensional representation of brain morphology"

| | | | | | | | |
|---|---|---|---|---|---|---|---|
| 036_S_4714 | 036_S_4715 | 036_S_4736 | 036_S_4740 | 036_S_4820 | 036_S_4878 | 036_S_4894 | 036_S_4899 |
| 036_S_5063 | 036_S_5112 | 036_S_5210 | 036_S_5248 | 036_S_5271 | 036_S_5283 | 037_S_4001 | 037_S_4015 |
| 037_S_4028 | 037_S_4030 | 037_S_4071 | 037_S_4146 | 037_S_4214 | 037_S_4302 | 037_S_4308 | 037_S_4381 |
| 037_S_4410 | 037_S_4432 | 037_S_4706 | 037_S_4750 | 037_S_4770 | 037_S_4879 | 037_S_5126 | 037_S_5162 |
| 037_S_5222 | 041_S_4004 | 041_S_4014 | 041_S_4037 | 041_S_4041 | 041_S_4051 | 041_S_4060 | 041_S_4138 |
| 041_S_4143 | 041_S_4200 | 041_S_4271 | 041_S_4427 | 041_S_4510 | 041_S_4513 | 041_S_4629 | 041_S_4720 |
| 041_S_4874 | 041_S_4876 | 041_S_4877 | 041_S_4974 | 041_S_4989 | 041_S_5026 | 041_S_5078 | 041_S_5082 |
| 041_S_5097 | 041_S_5100 | 041_S_5131 | 041_S_5141 | 041_S_5204 | 041_S_5244 | 041_S_5253 | 051_S_4929 |
| 051_S_4980 | 051_S_5285 | 051_S_5294 | 052_S_4626 | 052_S_4807 | 052_S_4885 | 052_S_4944 | 052_S_4945 |
| 052_S_4959 | 052_S_5062 | 053_S_4557 | 053_S_4578 | 053_S_4661 | 053_S_4813 | 053_S_5070 | 053_S_5202 |
| 053_S_5208 | 053_S_5272 | 053_S_5287 | 053_S_5296 | 057_S_4888 | 057_S_4897 | 057_S_4909 | 057_S_5199 |
| 057_S_5292 | 067_S_4054 | 067_S_4072 | 067_S_4184 | 067_S_4212 | 067_S_4310 | 067_S_4728 | 067_S_4767 |
| 067_S_4782 | 067_S_4918 | 067_S_5159 | 067_S_5160 | 067_S_5205 | 068_S_4061 | 068_S_4067 | 068_S_4134 |
| 068_S_4174 | 068_S_4217 | 068_S_4274 | 068_S_4332 | 068_S_4340 | 068_S_4424 | 068_S_4431 | 068_S_4859 |
| 068_S_4914 | 068_S_4968 | 068_S_5146 | 068_S_5206 | 070_S_4692 | 070_S_4708 | 070_S_4719 | 070_S_4793 |
| 070_S_4856 | 070_S_5040 | 072_S_4007 | 072_S_4057 | 072_S_4063 | 072_S_4102 | 072_S_4103 | 072_S_4131 |
| 072_S_4206 | 072_S_4226 | 072_S_4383 | 072_S_4390 | 072_S_4391 | 072_S_4394 | 072_S_4445 | 072_S_4462 |
| 072_S_4465 | 072_S_4522 | 072_S_4539 | 072_S_4610 | 072_S_4613 | 072_S_4694 | 072_S_4769 | 072_S_4871 |
| 072_S_4941 | 072_S_5207 | 073_S_4155 | 073_S_4216 | 073_S_4259 | 073_S_4300 | 073_S_4311 | 073_S_4312 |
| 073_S_4360 | 073_S_4382 | 073_S_4393 | 073_S_4403 | 073_S_4443 | 073_S_4540 | 073_S_4552 | 073_S_4559 |
| 073_S_4614 | 073_S_4739 | 073_S_4762 | 073_S_4795 | 073_S_4825 | 073_S_4853 | 073_S_4986 | 073_S_5016 |
| 073_S_5023 | 073_S_5090 | 073_S_5167 | 073_S_5227 | 082_S_4090 | 082_S_4208 | 082_S_4224 | 082_S_4244 |
| 082_S_4339 | 082_S_4428 | 082_S_5014 | 082_S_5029 | 082_S_5184 | 082_S_5278 | 082_S_5279 | 082_S_5282 |
| 094_S_4089 | 094_S_4162 | 094_S_4234 | 094_S_4282 | 094_S_4434 | 094_S_4459 | 094_S_4503 | 094_S_4560 |
| 094_S_4630 | 094_S_4649 | 094_S_4737 | 094_S_4858 | 098_S_4003 | 098_S_4018 | 098_S_4050 | 098_S_4059 |
| 098_S_4201 | 098_S_4215 | 098_S_4275 | 098_S_4506 | 099_S_4022 | 099_S_4076 | 099_S_4086 | 099_S_4104 |
| 099_S_4157 | 099_S_4202 | 099_S_4205 | 099_S_4463 | 099_S_4475 | 099_S_4480 | 099_S_4498 | 099_S_4565 |
| 099_S_4994 | 100_S_4469 | 100_S_4512 | 100_S_4556 | 100_S_5075 | 100_S_5091 | 100_S_5096 | 100_S_5102 |
| 100_S_5106 | 100_S_5280 | 109_S_4260 | 109_S_4380 | 109_S_4455 | 109_S_4499 | 109_S_4531 | 109_S_4594 |
| 114_S_4379 | 114_S_4404 | 114_S_5047 | 114_S_5234 | 116_S_4010 | 116_S_4043 | 116_S_4092 | 116_S_4167 |
| 116_S_4175 | 116_S_4195 | 116_S_4199 | 116_S_4209 | 116_S_4338 | 116_S_4453 | 116_S_4483 | 116_S_4625 |
| 116_S_4635 | 116_S_4855 | 116_S_4898 | 123_S_4096 | 123_S_4127 | 123_S_4170 | 123_S_4362 | 123_S_4526 |
| 123_S_4780 | 123_S_4806 | 123_S_4904 | 126_S_4458 | 126_S_4494 | 126_S_4507 | 126_S_4514 | 126_S_4675 |
| 126_S_4686 | 126_S_4712 | 126_S_4743 | 126_S_4891 | 126_S_4896 | 126_S_5214 | 126_S_5243 | 127_S_4148 |
| 127_S_4197 | 127_S_4198 | 127_S_4210 | 127_S_4240 | 127_S_4301 | 127_S_4500 | 127_S_4604 | 127_S_4624 |
| 127_S_4645 | 127_S_4765 | 127_S_4843 | 127_S_4844 | 127_S_4928 | 127_S_4940 | 127_S_4992 | 127_S_5028 |
| 127_S_5056 | 127_S_5058 | 127_S_5067 | 127_S_5095 | 127_S_5132 | 127_S_5185 | 127_S_5200 | 127_S_5218 |
| 127_S_5228 | 127_S_5266 | 128_S_4553 | 128_S_4571 | 128_S_4586 | 128_S_4599 | 128_S_4603 | 128_S_4607 |
| 128_S_4609 | 128_S_4636 | 128_S_4653 | 128_S_4671 | 128_S_4742 | 128_S_4745 | 128_S_4772 | 128_S_4774 |
| 128_S_4792 | 128_S_4832 | 128_S_4842 | 128_S_5066 | 128_S_5123 | 129_S_4073 | 129_S_4220 | 129_S_4287 |
| 129_S_4369 | 129_S_4371 | 129_S_4396 | 129_S_4422 | 130_S_4250 | 130_S_4294 | 130_S_4343 | 130_S_4352 |
| 130_S_4405 | 130_S_4415 | 130_S_4417 | 130_S_4468 | 130_S_4542 | 130_S_4589 | 130_S_4605 | 130_S_4641 |
| 130_S_4660 | 130_S_4730 | 130_S_4817 | 130_S_4883 | 130_S_4925 | 130_S_4971 | 130_S_4982 | 130_S_4984 |
| 130_S_4990 | 130_S_4997 | 130_S_5006 | 130_S_5059 | 130_S_5142 | 130_S_5175 | 130_S_5258 | 131_S_5138 |
| 131_S_5148 | 135_S_4281 | 135_S_4309 | 135_S_4356 | 135_S_4406 | 135_S_4446 | 135_S_4489 | 135_S_4566 |
| 135_S_4598 | 135_S_4657 | 135_S_4676 | 135_S_4689 | 135_S_4722 | 135_S_4723 | 135_S_4863 | 135_S_4954 |
| 135_S_5015 | 135_S_5113 | 135_S_5269 | 135_S_5273 | 135_S_5275 | 136_S_4189 | 136_S_4269 | 136_S_4408 |
| 136_S_4433 | 136_S_4517 | 136_S_4993 | 137_S_4211 | 137_S_4258 | 137_S_4299 | 137_S_4303 | 137_S_4331 |
| 137_S_4351 | 137_S_4466 | 137_S_4482 | 137_S_4520 | 137_S_4536 | 137_S_4587 | 137_S_4596 | 137_S_4623 |
| 137_S_4631 | 137_S_4632 | 137_S_4672 | 137_S_4678 | 137_S_4756 | 137_S_4815 | 137_S_4816 | 137_S_4852 |
| 137_S_4862 | 141_S_4053 | 141_S_4160 | 141_S_4232 | 141_S_4423 | 141_S_4426 | 141_S_4438 | 141_S_4456 |
| 141_S_4711 | 141_S_4803 | 141_S_4907 | 141_S_4976 | 153_S_4077 | 153_S_4125 | 153_S_4133 | 153_S_4139 |
| 153_S_4151 | 153_S_4159 | 153_S_4172 | 153_S_4297 | 153_S_4372 | 153_S_4621 | 153_S_4838 | 153_S_5261 |

**Supplementary Table S9.** List of ADNI subjects' ID used in this study (continuation).
