## Supplementary Table 10 for "Predicting cognitive decline in a low-dimensional representation of brain morphology"

| | | | | | | | |
|---|---|---|---|---|---|---|---|
| 153_S_5267 | 941_S_4036 | 941_S_4066 | 941_S_4100 | 941_S_4187 | 941_S_4255 | 941_S_4292 | 941_S_4365 |
| 941_S_4376 | 941_S_4377 | 941_S_4420 | 941_S_4764 | 941_S_5193 | 070_S_4798 | 022_S_4196 | 035_S_6739 |
| 037_S_6115 | 037_S_6125 | 037_S_6222 | 005_S_6084 | 007_S_6120 | 002_S_6103 | 019_S_6635 | 019_S_6668 |
| 011_S_6465 | 109_S_6220 | 099_S_6097 | 067_S_6117 | 114_S_6113 | 130_S_6027 | 130_S_6105 | 130_S_6111 |
| 135_S_6110 | 123_S_6118 | 127_S_6024 | 341_S_6605 | 141_S_6116 | 168_S_6131 | 168_S_6413 | 941_S_6471 |

**Supplementary Table S10.** List of ADNI subjects' ID used in this study (continuation).
