## Supplementary Figure 1 for "Predicting cognitive decline in a low-dimensional representation of brain morphology"

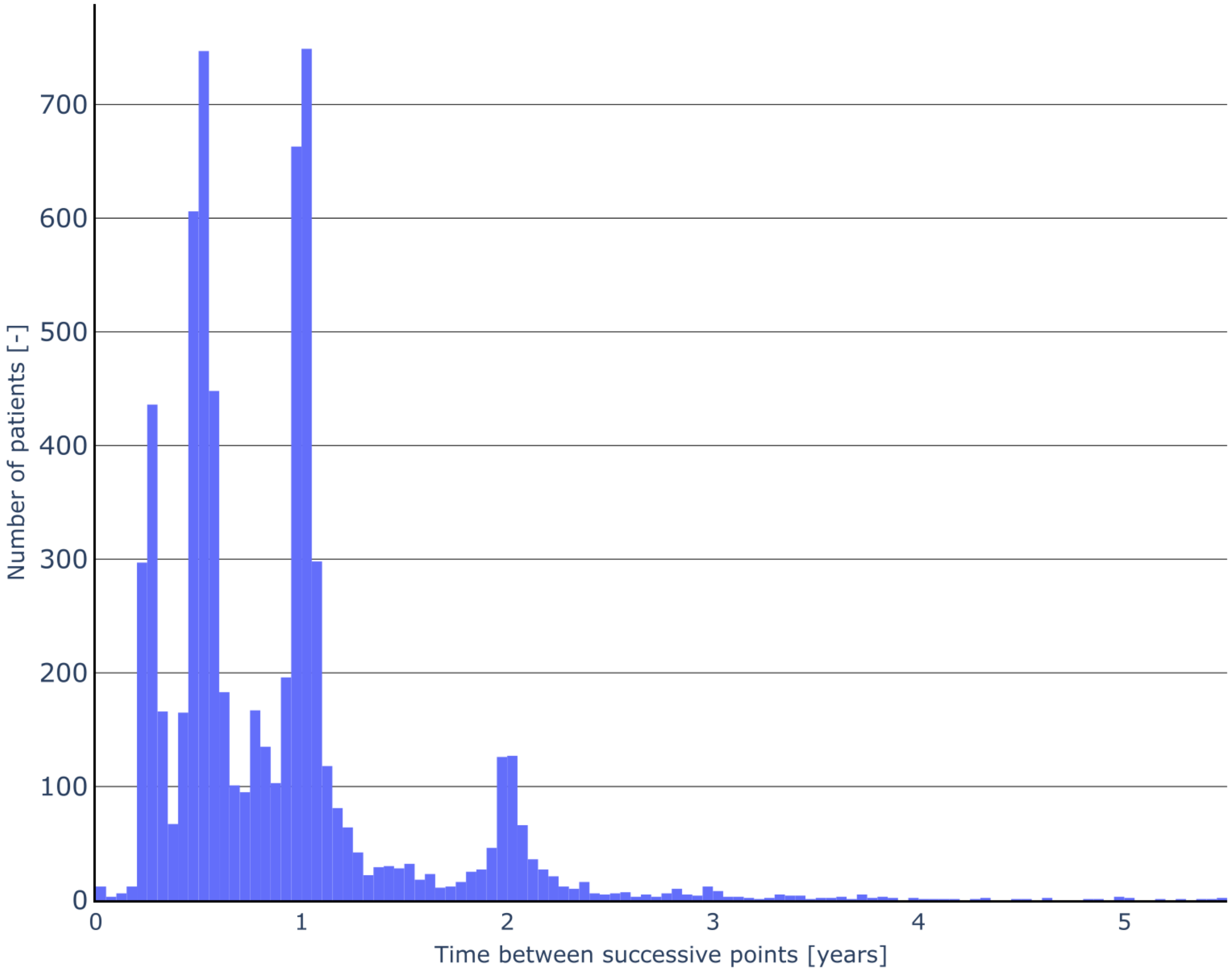


**Supplementary Figure S1.** Time step distribution for the ADNI database. The distribution of time between successive subjects' scan, for ADNI database.
