## Supplementary Figure 2 for "Predicting cognitive decline in a low-dimensional representation of brain morphology"

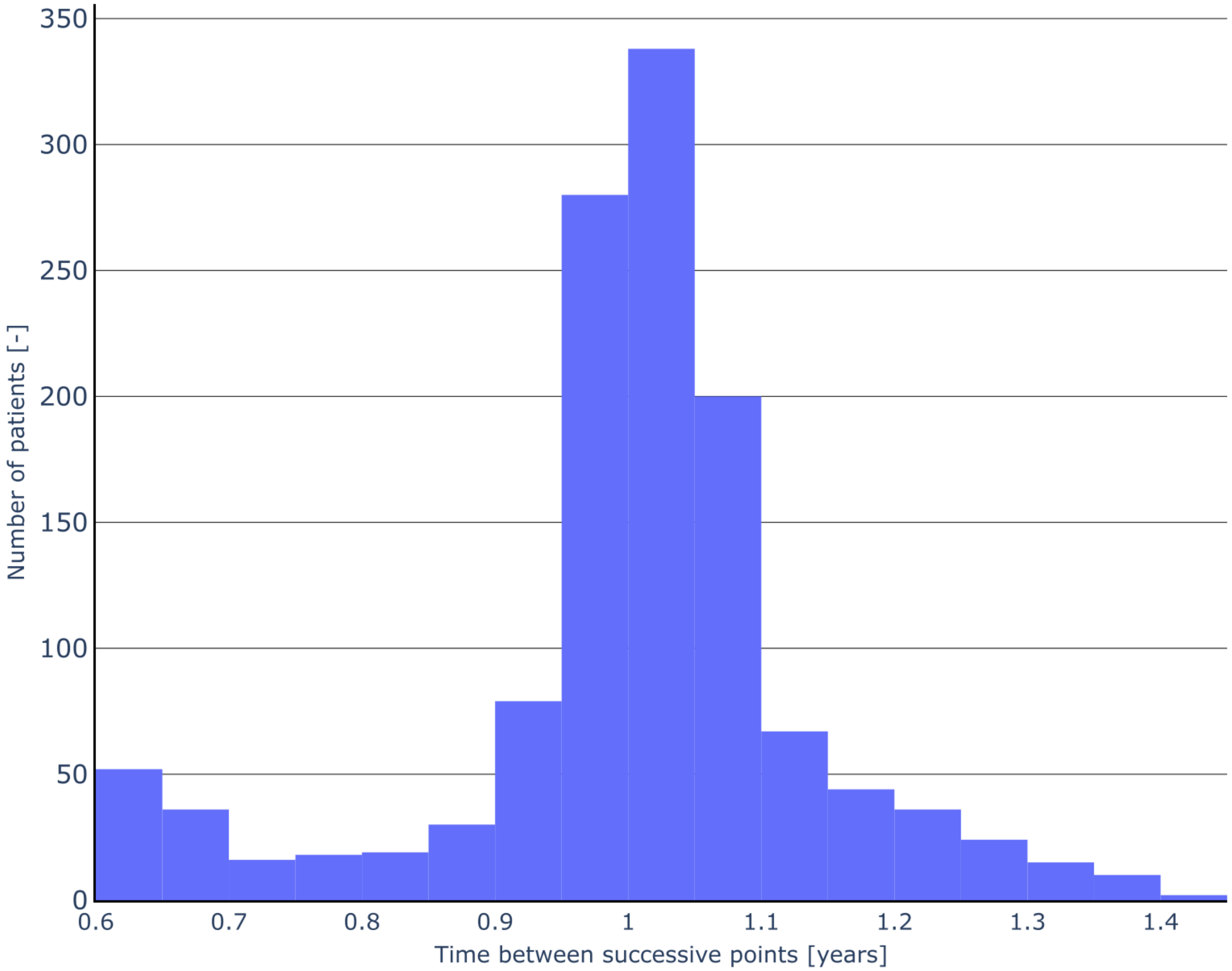


**Supplementary Figure S2.** Time step distribution for the standardized ADNI database. The distribution of time between successive subjects' scan, for ADNI database after having standardized this $\Delta t$. The standardization consist of removing data points so that the time between scan is always $1 \pm 0.4$ year. Subjects with one time point are also removed.
