## Supplementary figures and images for "Predicting cognitive decline in a low-dimensional representation of brain morphology"

### Supplementary Figure 3

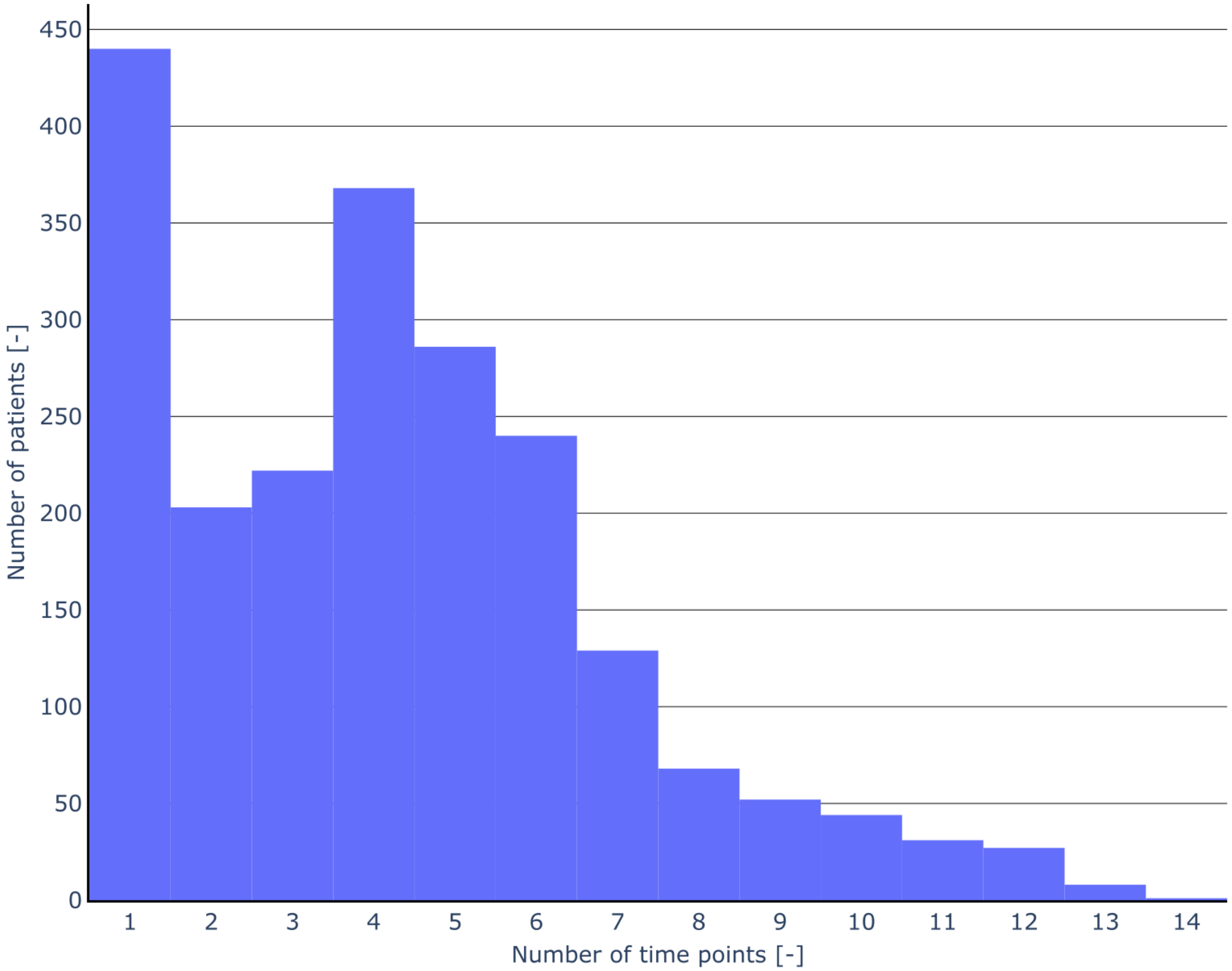


**Supplementary Figure S3.** The distribution of the number of scans in ADNI.
