## Supplementary Figure 4 for "Predicting cognitive decline in a low-dimensional representation of brain morphology"

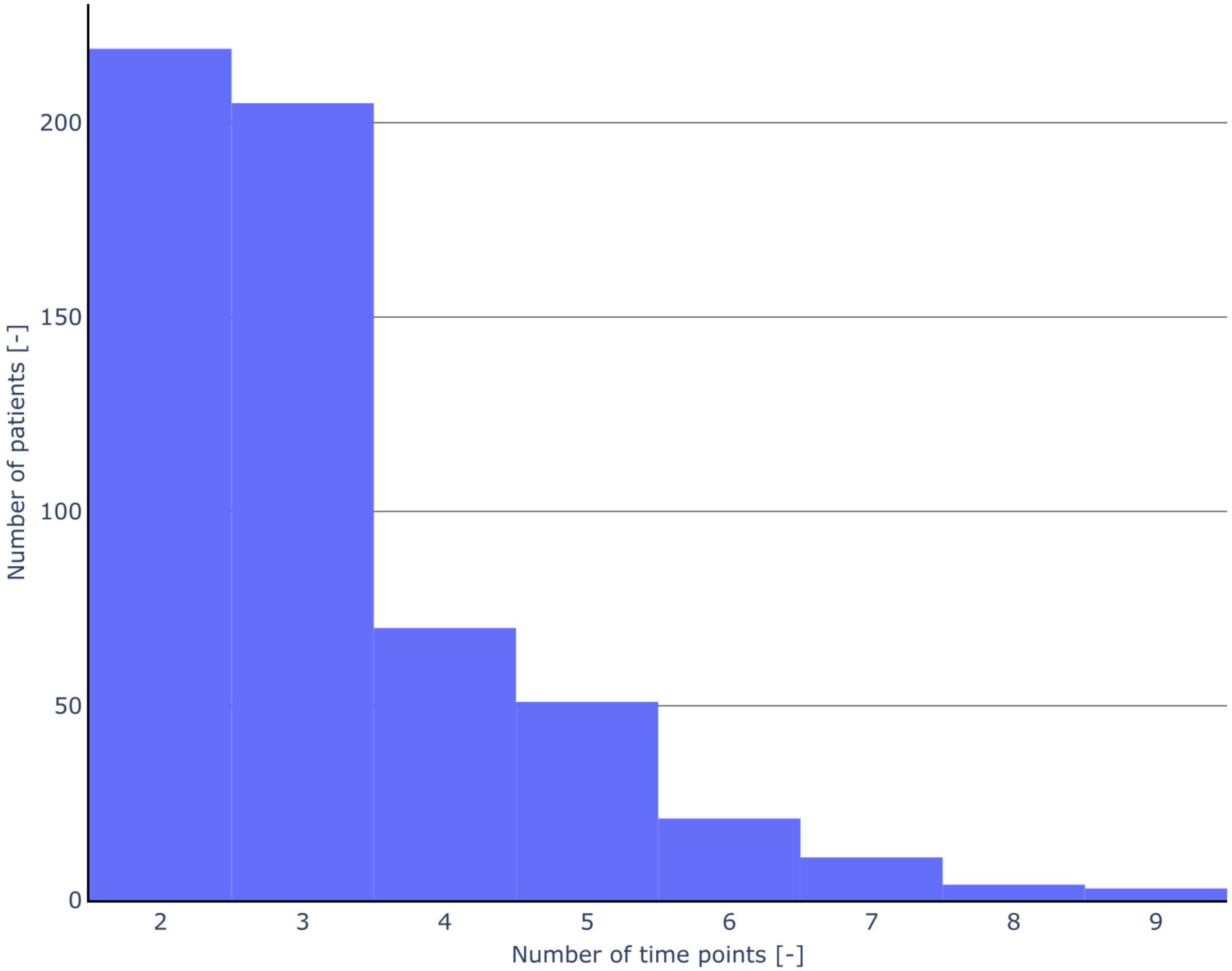


**Supplementary Figure S4.** The distribution of the number of scans in ADNI after standardization. Refer to Supplementary Figure S2 for a description of the standardization
