## Supplementary Figure 5 for "Predicting cognitive decline in a low-dimensional representation of brain morphology"

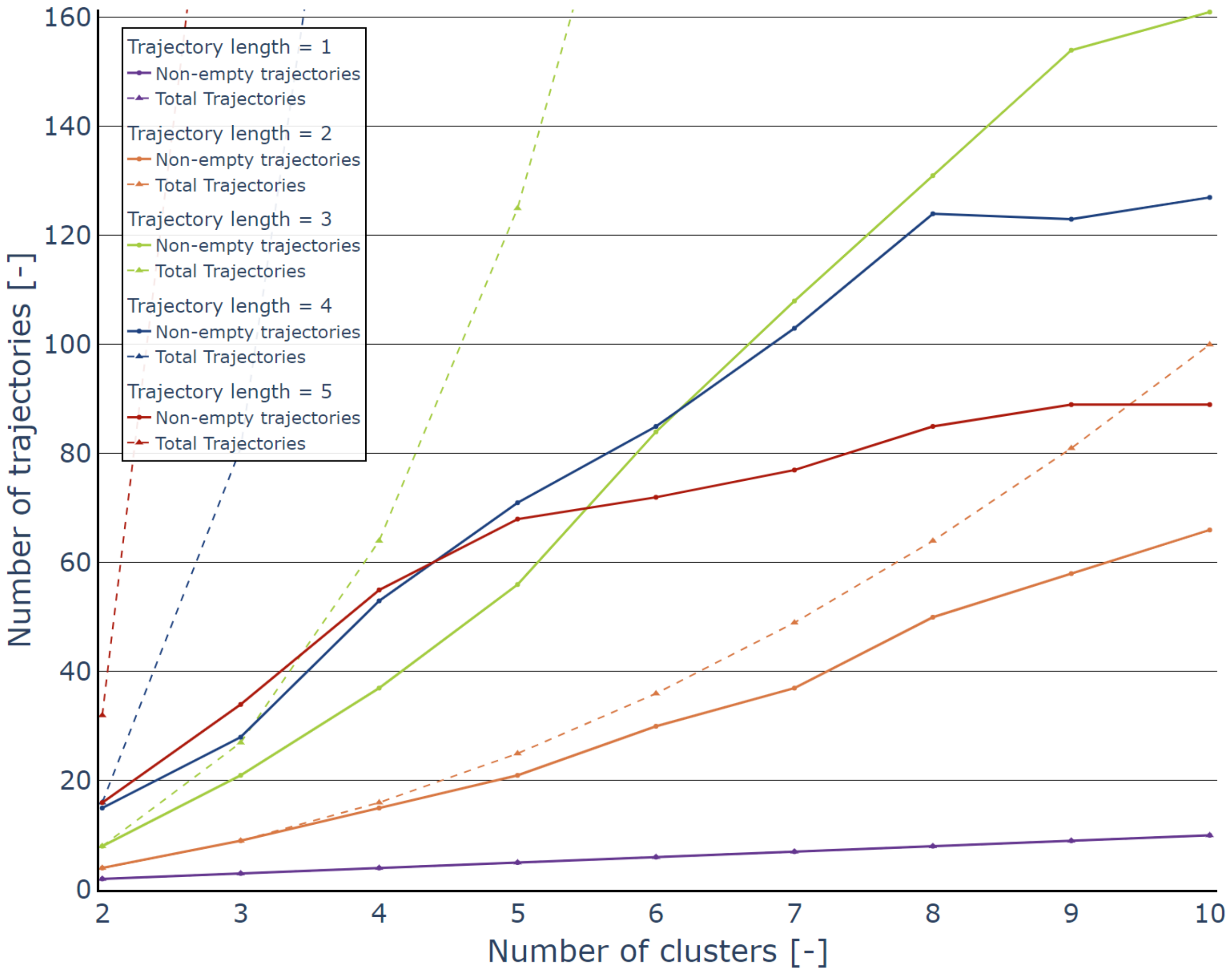


**Supplementary Figure S5.** The population in each trajectory by number of clusters ($C$). These values were obtained from the embedding with `n_neighbors`=20 and `min_dist`=0. The trajectory lengths ($l$) are colorcoded. The full lines are the number of trajectories that host at least one subject, by $C$, while the dashed lines are the total number of trajectories available as a function of $C$, also given by $C^l$. It is possible to see that from a trajectory length of 4, increasing the value reduces the number of trajectories that can be use. For a length of 4, the number of trajectories falls below the length of 3 after 6 clusters and the same happens for the curve with trajectory length of 5 after 5 clusters.
