## Supplementary Figure 6 for "Predicting cognitive decline in a low-dimensional representation of brain morphology"

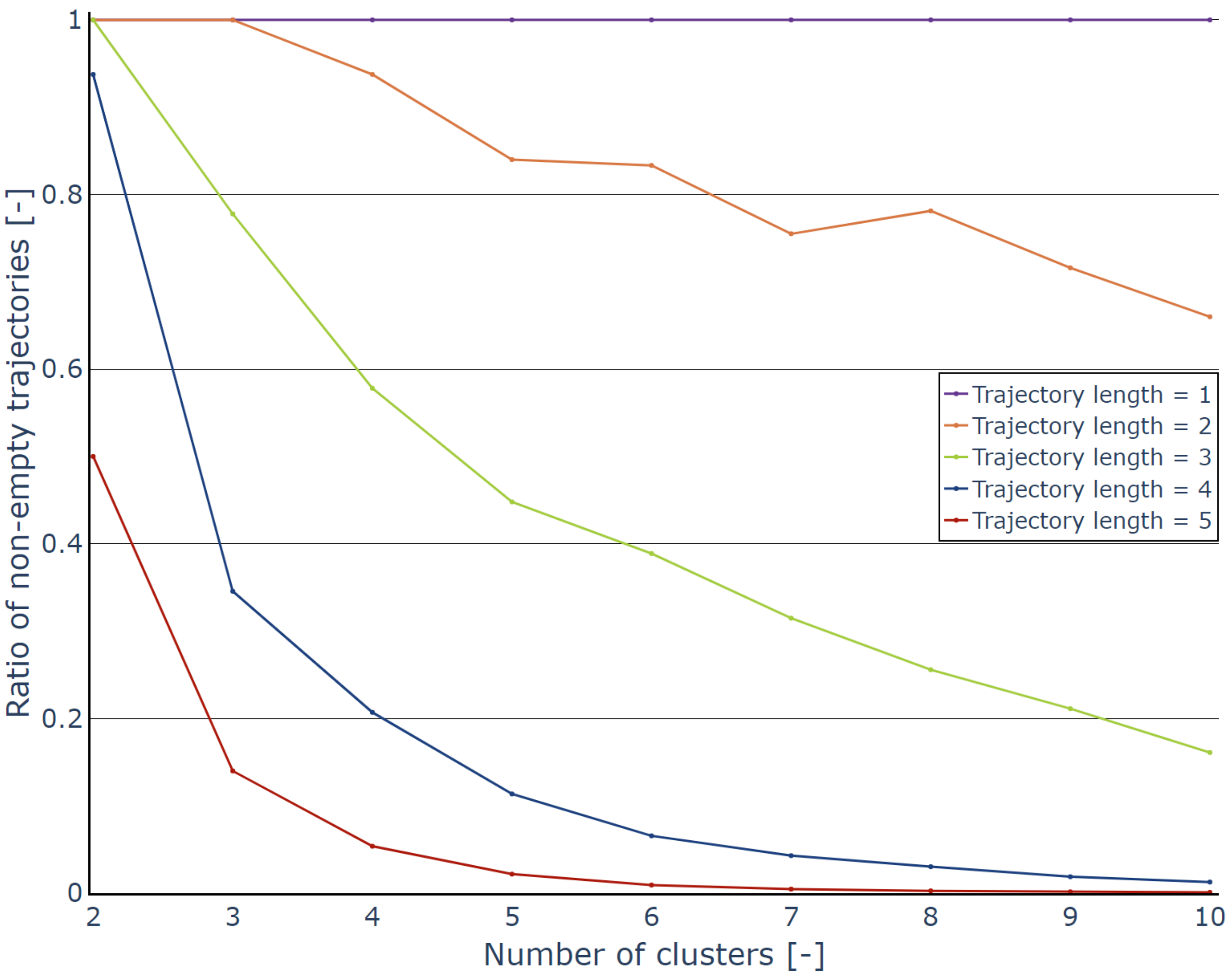


**Supplementary Figure S6.** The ratio of trajectories with at least one subject as a function of the number of clusters ($C$). These values were obtained from the embedding with `n_neighbors`=20 and `min_dist`=0. The trajectory lengths ($l$) are colorcoded. Similarly to Supplementary Figure S5, this figure showcases that too high $l$ and $C$ has diminishing returns. For $l \geq 4$ the ratio falls quickly under 20% and reaches $\approx$ 1% soon after. With the ADNI data, a maximum $l$ of 3 seems to minimise the number of unpopulated trajectories
